# The importance of sustained compliance with physical distancing during COVID-19 vaccination rollout

**DOI:** 10.1101/2021.09.22.21263944

**Authors:** Alexandra Teslya, Ganna Rozhnova, Thi Mui Pham, Daphne A van Wees, Hendrik Nunner, Noortje G Godijk, Martin Bootsma, Mirjam E Kretzschmar

## Abstract

Mass vaccination campaigns against SARS-CoV-2 are ongoing in many countries with increasing vaccination coverage enabling relaxation of lockdowns. Vaccination rollout is frequently supplemented with advisory from public health authorities for continuation of physical distancing measures. Compliance with these measures is waning while more transmissible virus variants such as Alpha (B.1.1.7) and Delta (B.1.617.2) have emerged. In this work, we considered a population where the waning of compliance depends on vaccine coverage. We used a SARS-CoV-2 transmission model which captures the feedback between compliance, infection incidence, and vaccination coverage to investigate factors that contribute to the increase of the prevalence of infection during the initial stages of the vaccination rollout as compared to no vaccination scenario. We analysed how the vaccine uptake rate affects cumulative numbers of new infections three and six months after the start of vaccination. Our results suggest that the combination of fast waning compliance in non-vaccinated population, low compliance in vaccinated population and more transmissible virus variants may result in a higher cumulative number of new infections than in a situation without vaccination. These adverse effects can be alleviated if vaccinated individuals do not revert to pre-pandemic contact rates, and if non-vaccinated individuals remain compliant with physical distancing measures. Both require convincing, clear and appropriately targeted communication strategies by public health authorities.

**Significance Statement:** SARS-CoV-2 vaccination campaigns are in progress in many countries around the world. As the vaccination coverage increases, the compliance with physical distancing measures aimed at reducing virus transmission may decline. Using a socio-epidemiological model we identify factors that are the drivers of increased transmission when SARS-CoV-2 prevalence is higher than the projected prevalence without vaccination. To maximize the benefits of vaccination campaigns, compliance in vaccinated and non-vaccinated groups should be targeted prioritizing one group over the other depending on the vaccination rate, the efficacy of vaccine in blocking the infection, and the circulating variant.

---

**M**ore than one year after the outbreak of COVID-19 was declared a pandemic by the World Health Organisation (1), the state of the pandemic in many countries around the world remained dire, with hospitalisations and death tolls mounting. Amid the second wave that started in fall 2020 (2–4), more transmissible (5–7) SARS-CoV-2 variants emerged (e.g., Alpha (B.1.1.7), Beta (B.1.351), and P.1 (Gamma) (8, 9), causing many countries to reinforce physical distancing measures in order to maintain healthcare capacities and to prevent deaths caused by COVID-19. Since then, an even more infectious virus variant, Delta, emerged, became dominant in Europe (10) and the US (11), and caused new pandemic waves. These events underscored that the physical distancing measures, while effective in significantly reducing SARS-CoV-2 transmission during the first wave (12–16), alone are not sufficient to limit SARS-CoV-2 transmission and to eradicate the need for future lockdowns, and further measures such as rigorous vaccination campaigns are required.

Fortunately, on the eve of spread of the Alpha variant in Europe, COVID-19 vaccines developed by BioNTech/Pfizer, Moderna, AstraZeneca, and Johnson & Johnson (Janssen) (17) were approved by EMA, fuelling hopes for the end of lockdown periods and relaxation of physical distancing measures. Phase 3 randomised clinical trials reported vaccine efficacies for preventing laboratory-confirmed symptomatic SARS-CoV-2 infection of 92% for BioNTech/Pfizer (18), 94.1% for Moderna (19), and at least 62% for AstraZeneca (20). The data from the vaccination campaign in Israel supports the results of the randomised trials for BioNTech/Pfizer vaccine reporting that the effectiveness of this vaccine against symptomatic disease is 94% (21). On 29 March 2021 CDC released a report that the Pfizer/BioNTech and Moderna vaccines have 80% effectiveness in preventing COVID-19 following 14 days or more after the first dose, but before the second dose, and 90% effectiveness following 14 or more days since the second dose (22). These findings are consistent with earlier reports that the three vaccines (Pfizer/BioNTech, Moderna and AstraZeneca) have some effectiveness in blocking SARS-CoV-2 transmission (23–25). Studies based on data collected in Israel estimated that the BNT162b2 mRNA COVID-19 vaccine developed by Pfizer/BioNTech reduced the acquisition rate for asymptomatic SARS-CoV-2 infection by 80% (26) up to 95% (27). Similarly high efficacy against infection acquisition were reported for the mRNA-1273 COVID-19 vaccine developed by Moderna, NIAID (28). For the adenovirus Ad26.COV2.S COVID-19 vaccine developed by Janssen Pharmaceutical Companies the efficacy in preventing infection with SARS-CoV-2 is reported to be 76% (29). These results were estimated from data collected between December 2020 and April 2021 in the USA. During this period, the original variant and the Alpha variant (B.1.1.7) were the dominant circulating variants.

Since then, a new, more infectious variant, Delta (B.1.617.2), has emerged and became dominant in many European countries (10) and the USA (11). A recent study based on data from Israel estimated a significant reduction of BNT162b2 efficacy for the Delta variant in preventing infection, which was 64% after two doses (30). This estimate was supported by another report based on the data in a highly vaccinated health system workforce of California San Diego Health (31).

The understanding of how the deployment of these vaccines can impact transmission is complicated by the emergence of these new variants (32–34). Thus, to reduce the death toll and the burden on healthcare system, as well as to slow down the appearance rate of antigenically relevant mutations that may escape protection conferred by existing vaccines, a swift and rigorous vaccination campaign seems of utmost importance.

Vaccination rollout, however, faces multiple challenges. Public health services may be confronted with structural and logistical obstacles (e.g., sufficient supply size, capacity to administer shots (35–37)) illustrated by diverging vaccination rates among different countries (38). Another factor that may affect vaccination rollout is vaccine acceptance (36) that varies greatly across countries from 23.6% in Kuwait to 97% in Ecuador (39, 40). Mass vaccination may also have undesirable consequences such as reducing compliance with physical distancing measures. For example, vaccinated people may increase their contact rate as they perceive COVID-19 to pose a lower risk for them. On the other hand, the non-vaccinated individuals may also become less compliant with the physical distancing measures, relying on decreased transmission due to the growing vaccination coverage. Thus, both vaccinated and non-vaccinated people may be less compliant with physical distancing measures due to factors such as lack of motivation or lack of knowledge about the necessity to maintain compliance post-vaccination. Hence, compliance may decline with increasing vaccination coverage. A number of modeling studies have shown that the feedback between the epidemic dynamics and human behaviour has an important role in the disease transmission (41–43). In an earlier modeling study (43), we showed that relaxation of compliance with physical distancing measures beyond a threshold may cause a significant increase in new infections and hospitalisations. This concern is especially relevant at the start of the vaccination campaign, when vaccination coverage is still low.

We developed a socio-epidemiological model (Figure -1) for SARS-CoV-2 transmission to investigate the effects of waning of compliance with physical distancing measures on the dynamics of SARS-CoV-2 transmission as vaccine is rolled out in the population. The transmission dynamics is modelled through a susceptible-exposed-infectious-recovered (SEIR) framework. The vaccine works as all-or-nothing conferring perfect protection to a fraction of susceptible individuals who receive it. The vaccine delivered to individuals in other disease stages has no effect.

**Fig. 1.**
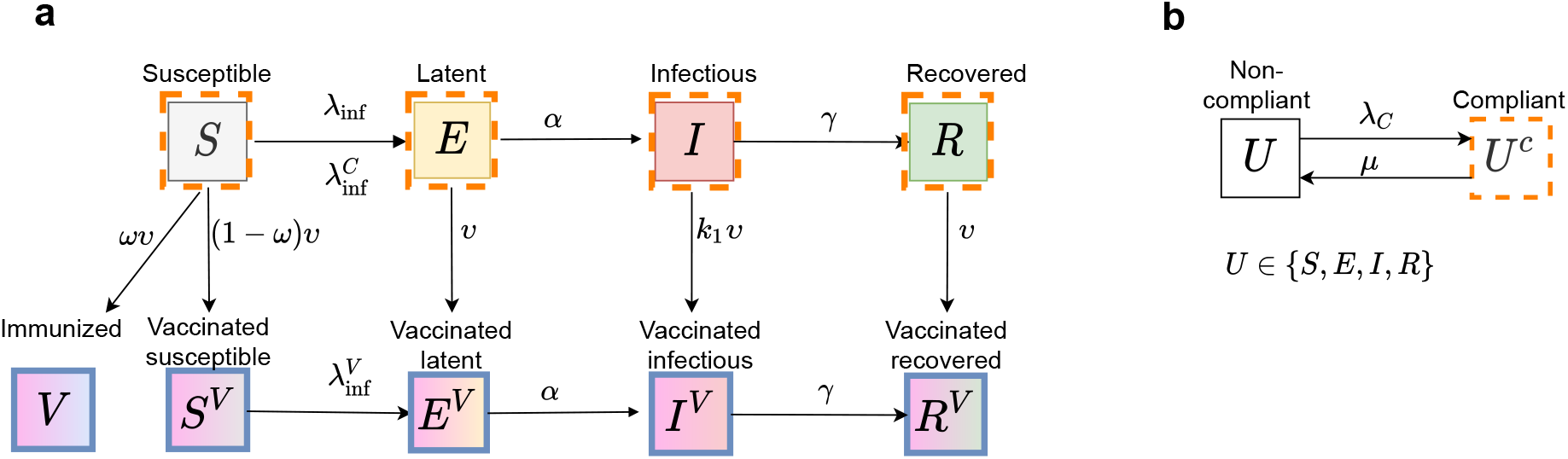
Flow diagram of the infection transmission dynamics coupled with compliance and vaccination processes. **a** Flow diagram of infection transmission and vaccination rollout, **b** Flow diagram of acquisition and loss of compliance. Solid-colored rectangles denote non-vaccinated compartments; solid-bordered rectangles denote non-compliant compartments; orange dashed-bordered rectangles denote compliant compartments; gradient-colored rectangles denote vaccinated compartments. Susceptible individuals (*S, S*^*C*^, and *S*^*V*^) become exposed (*E, E*^*C*^, and *E*^*V*^, respectively) with rates 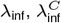, and 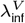 through contact with infectious individuals (*I, I*^*C*^, and *I*^*V*^). Exposed individuals become infectious (*I, I*^*C*^, and *I*^*V*^, respectively) at rate *α*. Infectious individuals recover (*R, R*^*C*^, and *R*^*V*^) at rate *γ*. Compliance is gained with rate *λ*^*C*^ and lost with rate *µ*. Individuals in any state of infection or compliance can get vaccinated. A proportion *ω* of susceptible individuals *S*, who were vaccinated are fully protected, *V*. Individuals who were vaccinated, but did not obtain protection, are denoted by *S*^*V*^, *E*^*V*^, *I*^*V*^ and *R*^*V*^ and are epidemiologically indistinguishable from their non-vaccinated counterparts.

We assume that the vaccination rollout takes place during a government-imposed lockdown, whereupon many public venues are closed or operate at a reduced capacity, thus limiting the average number of contacts. Additionally, the government may issue a set of recommendations with respect to physical distancing. Compliance with these recommendations is captured by a reduction in the daily number of contacts relative to the pre-pandemic level of contacts. The non-vaccinated population is divided into individuals who can be more compliant (henceforth referred to as “compliant”) and less compliant (“non-compliant”) to measures. The reduction in contacts is larger for compliant and smaller for non-compliant populations. On the other hand, we assume that vaccinated individuals perceive themselves protected from COVID-19 and therefore, are no longer compelled to comply with physical distancing measures. Thus they are not affected by the compliance acquisition-loss process and increase their contact rate above that of non-compliant individuals, thereby returning to nearly pre-pandemic level of contacts. Non-vaccinated individuals can move between compliant and non-compliant modes, and the rates of moving depend on the state of the epidemic and on vaccination coverage. Specifically, more individuals become compliant with physical distancing measures as the incidence of SARS-CoV-2 infection cases increases and lose compliance faster as the proportion of vaccinated individuals grows (see Methods).

We considered a baseline scenario without vaccination and several vaccination scenarios. To observe the full spectrum of possible scenarios, we sampled vaccination rate on a wide range, which was based on the observations during the first six months of the vaccination rollout in European countries and Israel (38). Further, we considered scenarios for three types of SARS-CoV-2 variants. The first variant has the transmission potential of the original variant that was circulating in Europe prior to fall 2020. The second variant is a more transmissible, Alpha-like variant (B.1.1.7), that spread in many European countries during the winter of 2020/2021 and became dominant in the spring of 2021 (44). Finally, we also considered the dynamics of a “hyper-contagious” Delta-like variant (B.1.617.2), which, as of August 2021, became the dominant strain in Europe (10). We investigated the impact of compliance with physical distancing measures on the numbers of infected, vaccinated and compliant individuals over the course of the vaccination rollout. We also compared the cumulative numbers of new infections after three and six months into the vaccination programme to the numbers without vaccination. We tested the robustness of our findings to the values we chose for the initial conditions and parameters by performing multivariate sensitivity analyses (see Figure 5 and Supplementary materials). The values for initial conditions and parameters were sampled continuously.

Next, we considered the potential effects of two interventions aimed at improving compliance. The first intervention is targeted at people who have not been vaccinated yet and aims at keeping their compliance with physical distancing at the level of prior to vaccination rollout. The second intervention is targeted at people who have been vaccinated and aims at keeping their contact rates low. We also considered a combined intervention where both interventions are implemented simultaneously.

Finally, we considered the possibility that in the case of a sharp rise in prevalence which may occur due to the decline of compliance with physical distancing measures, the government may impose additional physical distancing rules to reduce SARS-CoV-2 transmission. To wit, in the Netherlands, following a sharp increase in the number of detected infections in June of 2021, the government imposed additional measures which aimed to reduce infection transmission and which were in effect for nearly a month (July 10, 2021 to August 13, 2021) (45). We have investigated outcomes of the combination of the vaccination rollout with a lockdown which initiates when the prevalence of infectious cases surpasses a threshold.

## Results

### Compliance and vaccination rollout

To model the transmission dynamics of each of the SARS-CoV-2 virus variants that we consider, the model was calibrated to the state of the epidemic and the level of compliance with physical distancing measures prior to the start of vaccination in the Netherlands in November 2020. The size of the population that recovered from SARS-CoV-2 infection was set based on seroprevalence data from the serological study in an age-stratified and regionally weighted representative sample of the Dutch population (46, 47). The estimated seroprevalence was 4% in June/July of 2020 (46) and increased to 14% in February 2021 (47). To account for the effects of the second wave until the start of vaccination (taken in the simulations to be November 2020) we fixed the recovered population at 8%. The proportion of compliant population was set at 65% using the study on behavioral measures and well-being conducted in the Netherlands by the National Institute for Public Health and the Environment (RIVM) (48). We have investigated the sensitivity of the outcomes that we collected to the assumed initial conditions. We have assumed that the baseline epidemiological dynamics when each one of the three variants circulate are identical, except for for the probability of transmission per contact. We have assumed an Alpha-like virus variant to be approximately 1.5 times more infectious than the original variant and that a Delta-like variant to be approximately 2 times more infectious than the original variant.

The vaccination rates were sampled on an interval. The lowest boundary of the interval is based on the data from the first six months of vaccination rollout in Belarus (38), one of the slower vaccinating countries in Europe, and would lead to 10% of the population to be vaccinated in the first six months of the vaccination rollout. The upper boundary of the vaccination rate interval is based on the vaccination rollout in Israel in the first six months of the vaccination campaign. Sustaining this rate would lead to almost 60% of the population to be vaccinated six months after the start of the vaccination rollout. Henceforth, these rates are referred to as “slow” and “fast”, respectively. Figure **2a** shows vaccination coverage during the first six months after the start of vaccination rollout for slow and fast vaccination. In our analyses, we considered a wide range of vaccine efficacies with respect to prevention acquisition of the infection, 55% to 95%. For some of the results in the main analysis we have fixed the vaccine efficacy to 60% and subsequently explored sensitivity of the outcomes to this parameter.

**Fig. 2.**
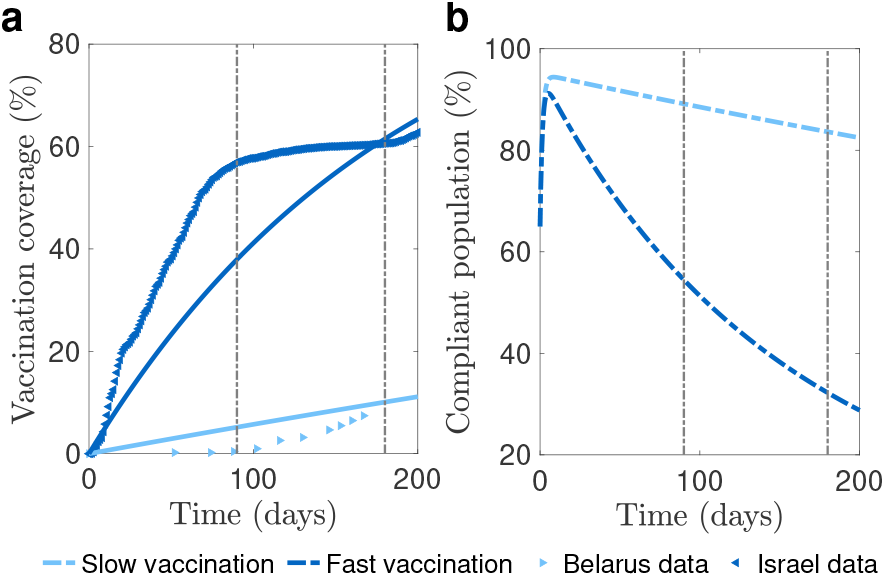
Vaccination coverage and proportion of compliant population during the vaccination rollout. **a** Increase in vaccination coverage for slow (light blue line) and fast (dark blue line) vaccination rates. Stars show data for Belarus (light blue) and Israel (dark blue) (38), respectively. **b** Decrease in the proportion of compliant population for slow and fast vaccination and a fixed incidence of infection (16,062 cases per day) observed in the Netherlands in the period used for the model calibration. Vertical brown lines mark three and six months since the start of vaccination.

We fixed the contact rate for compliant and non-compliant individuals such that the effective reproduction number for the original variant prior to vaccination rollout is 1.1, as estimated for the Netherlands in November 2020 (49). The effective reproduction number for the Alpha-like variant was 1.65, i.e. 50% higher than for the original variant (6). We have set the basic reproductive number for the Delta-like variant using the estimate of 4.92 (50), which makes it approximately 2 times more transmissible as the original variant. Therefore, the effective reproductive number for the Delta-like variant was approximately equal to 2.2 at the start of the vaccination rollout. The contact rate of vaccinated individuals was assumed to be close to the pre-pandemic rate and 1.5 times higher than the contact rate of non-compliant individuals (51). We explored sensitivity of the outcomes to this parameter.

In our model, individuals become compliant if there are infectious individuals in the population. The per capita rate of switching to the compliant state is proportional to the incidence of infectious cases (see Methods, Table 1). The rate of moving to the compliant state was fixed in the main analysis. The sensitivity analyses for this parameter are shown in the Supplementary materials. Furthermore, the compliance which has an intrinsic natural decay rate, wanes more rapidly as the vaccination coverage increases. The proportion of compliant population for a constant incidence of infection is shown in Figure 2**b** where we used slow and fast vaccination rates from Figure 2**a**. We used incidence of 16,062 cases per day, based on the number of infectious people in the Netherlands which was approximated by RIVM using hospital admissions and data from the Pienter Corona study (47) in the period used for the model calibration (4). For slow vaccination, three months after the start of vaccination, approximately 89% of the population is compliant with physical distancing measures and after six months, 84% is compliant. For fast vaccination, the compliant population decreases more rapidly, with only approximately 54% and 32% of individuals being compliant after three and six months, respectively.

**Table 1.** Summary of model parameters.

| Name | Description (unit) | Value* | Source |
| --- | --- | --- | --- |
| *Epidemiological parameters* |  |  |  |
| $R_0$ | Basic reproduction number, original variant | 2.5 | (60, 61) |
| $R_0^{\text{new}}$ | Basic reproduction number, Alpha (B.1.1.7)-like variant | 3.75 | (6, 7) |
| $R_0^{\text{new}}$ | Basic reproduction number, Delta (B.1.617.2)-like variant | 4.92 | (50) |
| $R_e$ | Effective reproduction number, original variant | 1.1 | Computed using the method in (62) |
| $\hat{c}$ | Average contact rate prior to the epidemic (individuals/day) | 14.9 | (51) |
| $\epsilon$ | Probability of transmission per contact, original variant | $2.4 \times 10^{-2}$ | From $R_0 = \hat{c}\epsilon/\gamma = 2.5$ |
| $\epsilon^{\text{Alpha}}$ | Probability of transmission per contact, Alpha-like variant | $3.6 \times 10^{-2}$ | From $R_0 = \hat{c}\epsilon^{\text{Alpha}}/\gamma = 3.75$ |
| $\epsilon^{\text{Delta}}$ | Probability of transmission per contact, Delta-like variant | $5.4 \times 10^{-2}$ | From $R_0 = \hat{c}\epsilon^{\text{Delta}}/\gamma = 5.63$ |
| $c$ | Average contact rate of non-compliant individuals starting November 16, 2020 (individuals/day) | 8.8 (0.5 – 15) | Obtained from solving $R_e(0) = 1.1$ |
| $r_1$ | Ratio between contact rates of compliant and non-compliant individuals | 0.34 (0.01 – 1) | Assumed, control parameter |
| $r_2$ | Ratio between contact rates of vaccinated and non-compliant individuals | 1.5 (1, 1.5) | Assumed, control parameter |
| $1/\alpha$ | Duration of latent period (days) | 4 (2-6) | (60, 61, 63) |
| $1/\gamma$ | Duration of infectious period (days) | 7 (5-9) | (64) |
| *Compliance parameters* |  |  |  |
| $\delta$ | Rate of moving to compliant state (1/day) | $4 \times 10^{-5}$ ($10^{-6} - 10^{-4}$) | Assumed, control parameter |
| $1/\mu_0$ | Duration of compliant state when there is no vaccination (days) | 30 (7 – 30) | Sensitivity analyses |
| $\mu_1$ | Parameter describing how loss of compliance increases depending on vaccination coverage (1/day) | 0, 0.3 | Sensitivity analyses |
| *Vaccination parameters* |  |  |  |
| $v$ | Vaccine uptake rate (1/day) | $(5, 60) \times 10^{-3}$ | Based on vaccination data in (38) |
| $\omega$ | Vaccine efficacy in conferring protection against becoming infected | 0.6 (0.55 – 0.95) | Based on estimates of efficacies in blocking SARS-CoV-2 infections for some of the existing vaccines (22, 24–27, 29–31, 59) for a variety of variants, control parameter |
| $k_1$ | Reduction factor in vaccination rate of infectious individuals | 1 | Assumed |
| *Lockdown parameters* |  |  |  |
|  | Threshold of infectious individuals for strengthening/relaxation of the lockdown (individuals) | 50 – 500 | Sensitivity analysis |
|  | Average contact rate during strengthened lockdown (individuals/day) | 3 | Sensitivity analysis |

\* Interval was used in sensitivity analyses.

The reason for the decline of compliance observed in Figure **2b** is two-fold. First, as the vaccination coverage increases, the compliance in the non-vaccinated population decreases. Moreover, the speed of this decrease depends on how fast vaccination is rolled out. Second, per our assumption, vaccinated people perceiving themselves protected from COVID-19, subsequently are no longer compelled to comply with physical distancing. These two processes translate into varying proportions of the compliant population depending on both the incidence of infection and vaccination coverage.

### Epidemic dynamics with vaccination

The model predicts that depending on the speed of the vaccination rollout and transmissibility of the virus variant, as a result of decreasing compliance with physical distancing measures, the prevalence of infected individuals in the presence of vaccination can be higher than the prevalence in a situation without vaccination (Figure 3). This effect is much more pronounced for the more transmissible Alpha-like and Delta-like variants than for the original variant (Figures **3a, 3b**, and **3c**). We quantify it as the difference in the cumulative number of new infections relative to the no-vaccination scenario level three and six months after the start of the vaccination rollout.

**Fig. 3.**
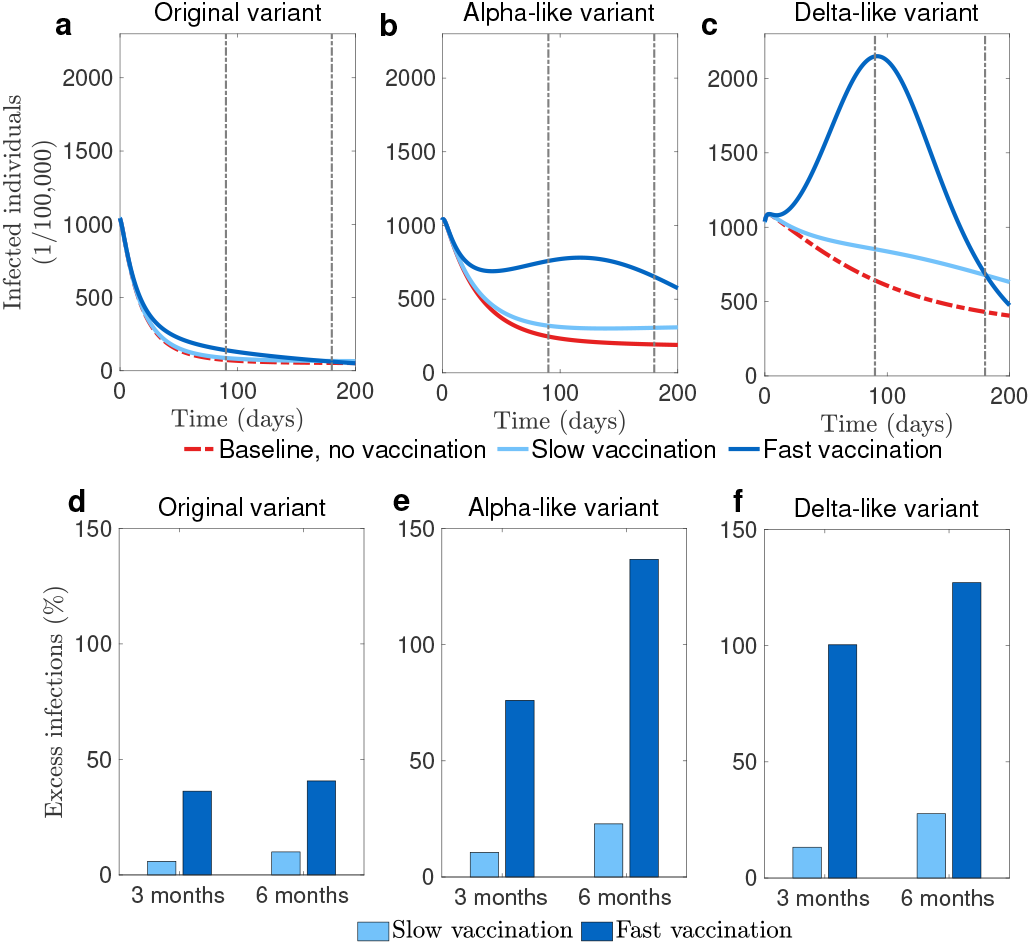
Epidemic dynamics with and without vaccination. **a** Prevalence of infected individuals versus time when the original variant circulates. **b** The same output when an Alpha-like variant circulates. **c** The same output when a Delta-like variant circulates. **d** Difference in the cumulative number of new infections relative to the no-vaccination scenario level for the original variant. **e** The same output when an Alpha-like variant circulates. **f** The same output when a Delta-like variant circulates. **d, e** and **f** show the difference in the cumulative number of new infections relative to the no-vaccination levels when respective variants circulate. In **a, b**, and **c**, vertical brown lines mark three and six months since the start of vaccination.

If the original variant is circulating (Figures 3**a** and 3**d**), vaccination can reduce the prevalence below the level of the no-vaccination scenario six months after the start of the campaign (Figure 3**a**). In our simulations, this happens when the vaccination rate is fast. When the vaccination rate is slow, as the result of the vaccination rollout, the prevalence eventually decreases below the level of the no-vaccination scenario, but it takes more than 600 days (result not shown). However, for both vaccination rates, due to the decline of compliance following the growing vaccination coverage, in the initial stages of the rollout, the transient prevalence can be higher than in the no-vaccination scenario. This difference in prevalence is higher for the fast vaccination rate than for the slow vaccination rate. Consequently, slow vaccination, if associated with waning of compliance during vaccine rollout, leads to a smaller excess of cumulative infections than fast vaccination at both three and six months time points (Figure 3**d**).

If a more transmissible variant is circulating (for example, an Alpha-like or a Delta-like) (Figures 3**b**, 3**c**, 3**e**, and 3**f**) decreased compliance with physical distancing measures can lead to an additional peak in prevalence (Figure 3**b**). Additionally, similar to the scenario with the original variant, vaccination can lead to an increase of cumulative number of new infections compared the no-vaccination scenario (Figures 3**e** and 3**f**). This occurs because waning of compliance coincides with an increased transmissibility of the virus. The period when the prevalence is higher as compared to the no-vaccination scenario lasts even longer than for the original variant (Figures 3**b** and 3**c**).

#### Contribution of vaccinated and non-vaccinated individuals to the attack rate

To understand the role of vaccinated individuals in the transmission dynamics observed in Figure 3, we calculated the proportion of infections occurring in the vaccinated population over time (Figure 4). The analyses show that in the case of slow vaccine uptake (Figures 4**a**, 4**b**, and 4**c**) vaccinated individuals comprise a small proportion of the infected population even at the end of the six months of the vaccination campaign. Therefore, the increased prevalence among non-vaccinated can be attributed to the decrease of their compliance with physical distancing measures. In the case of fast vaccine uptake, the model predicts that a proportion of infections among vaccinated individuals is higher. Moreover, for a “hyper-contagious” strain, similar to the Delta variant, and a vaccine with relatively low efficacy (60% in our main analysis), more than a third of infections are expected to be in the vaccinated population. Thus, the observed rise in the prevalence is in part due to the increased contact rate of susceptible vaccinated individuals. These findings suggest that for slow vaccination the risk of severe disease and death in the population is hardly lowered, while for fast vaccination a significant proportion of the infected individuals will be protected against severe disease, even if the incidence of cases is high.

**Fig. 4.**
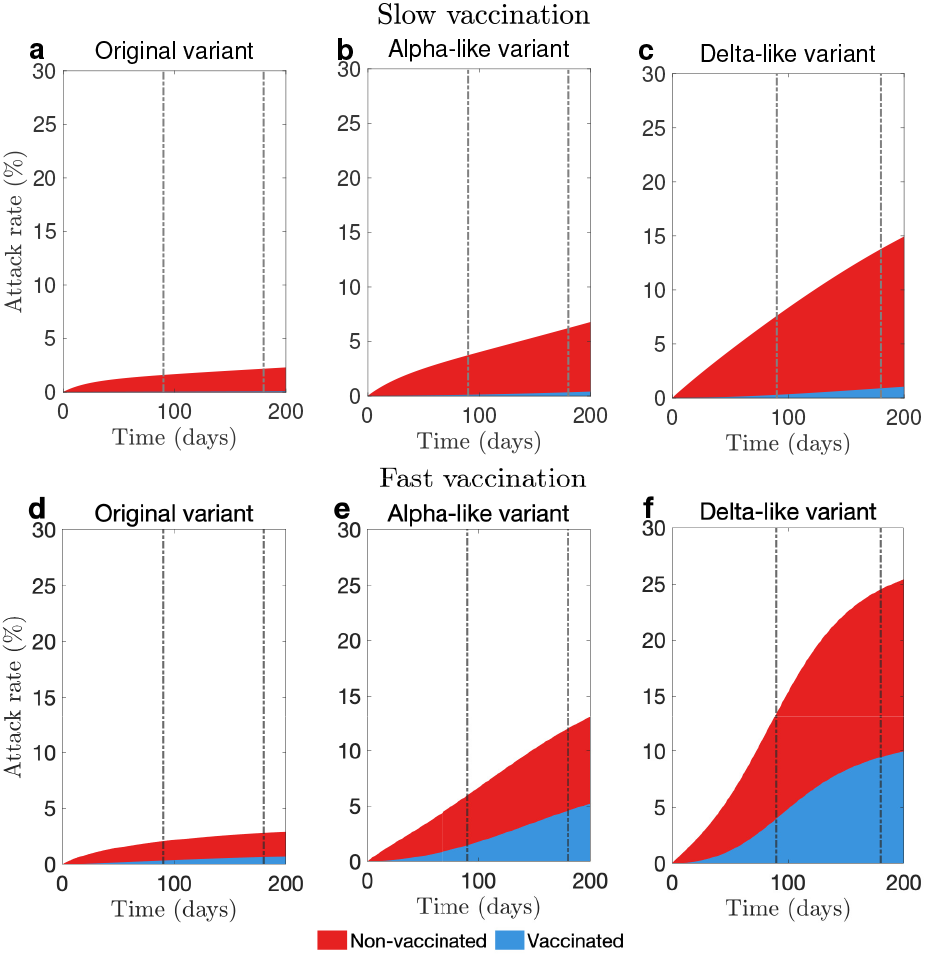
Contribution of vaccinated and non-vaccinated individuals to attack rate during the vaccination rollout. **a, b**, and **c** show attack rates versus time given the slow vaccine uptake rate. **d, e**, and **f** show attack rates versus time given the fast vaccine uptake rate. **a** and **d** show these quantities for the original variant, **b** and **e** for an Alpha-like variant, **c** and **f** for a Delta-like variant. Vertical brown lines mark three and six months since the start of the vaccination campaign. Attack rate is the proportion of the population that has been infected until a given time. We adjusted the attack rate so that it describes only new infections that appeared during the time interval that we considered.

#### Sensitivity of the vaccination rollout outcomes to vaccine efficacy and vaccine uptake rate

Finally, we investigated the effect of vaccine efficacy and vaccine uptake rate on the excess of the cumulative number of new infections as compared to the no-vaccination scenario three and six months after the start of vaccination rollout (Figures 5**a**, 5**b** and Figures **1a, 1b**, **2a**, and **2b** in Supplementary materials). In all panels, in the region above the magenta curve vaccination rollout yields improvement over the no-vaccination scenario, i.e. the cumulative number of new infections is lower. Importantly, the slower is the vaccination rollout, i.e. the lower is the vaccination coverage after three months of the vaccination rollout, the higher the vaccine efficacy needs to be to avoid an increase of cumulative number of new infections as compared to the no-vaccination scenario. This is a consequence of fast loss of compliance with physical distancing measures as the vaccination coverage grows. Vice-versa, depending on the vaccine efficacy, the speed of the rollout can cause increase or decrease of cumulative number of new infections. If the efficacy is low and the rollout is fast, then initially the cumulative number of new infections is expected to be higher than for the no-vaccination scenario. Moreover, the combination of fast vaccine uptake and low vaccine efficacy is predicted to cause the largest increase in the cumulative number of new infections as compared to the no-vaccination scenario. This happens due to the combined effect of quickly growing vaccination coverage which affects compliance with physical distancing measures in the non-vaccinated population and of increased contact rates of the vaccinated individuals who while potentially protected from the severe disease can still acquire and transmit the infection. However, if the vaccine efficacy is high, given a fast vaccination rate, we expect that the cumulative number of new infections to fall below the level of the no-vaccination scenario. The decrease in the number increases as the vaccination rate increases. We observe that for all variants considered (see Figures 5**a**, 5**b** and Figures **1a, 1b**, **2a**, and **2b** in Supplementary materials), the minimal vaccine efficacy where the cumulative number of new infections decreases over the no-vaccination scenario decreases with time since the start of the vaccination rollout.

**Fig. 5.**
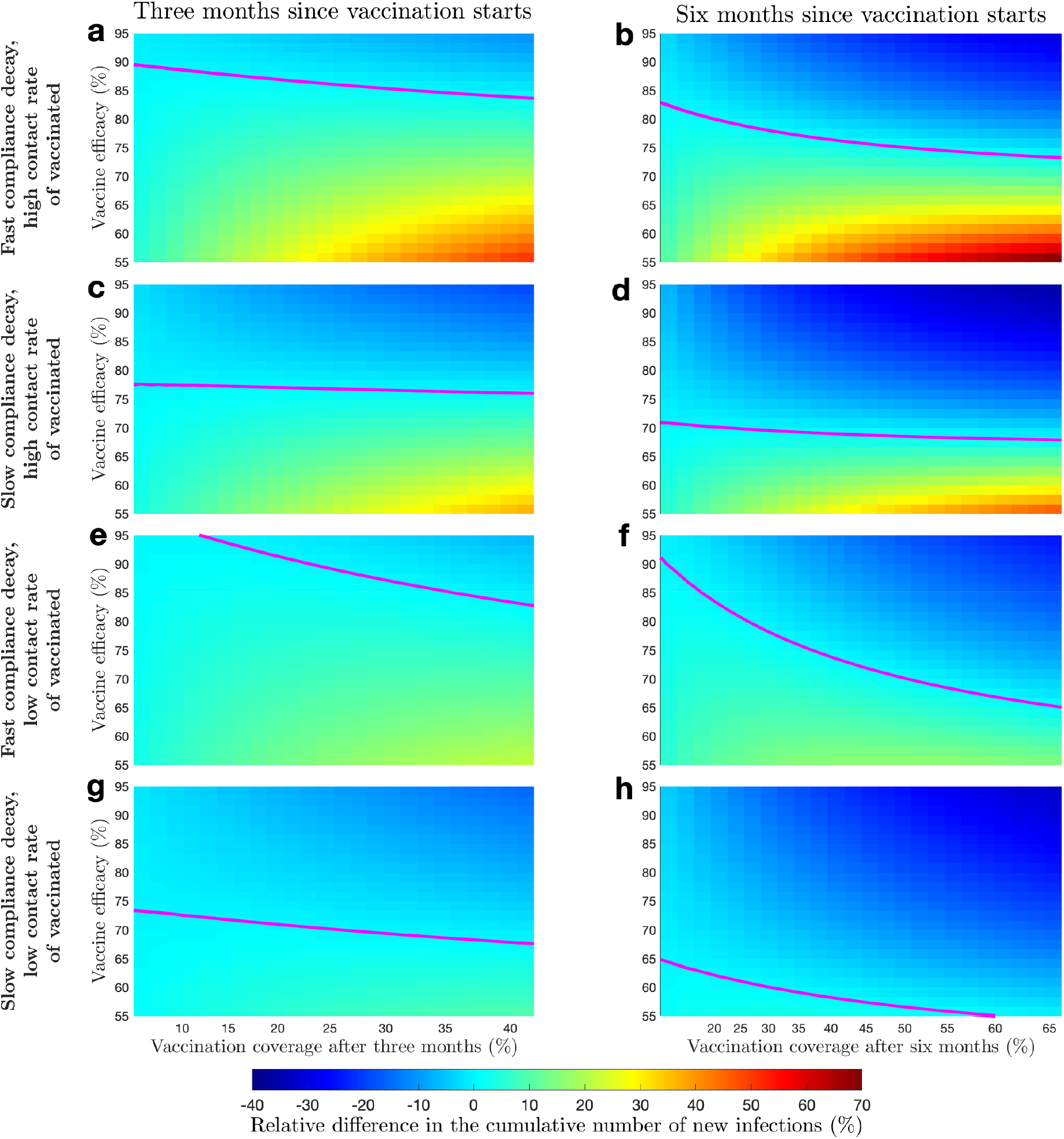
Epidemic dynamics with and without interventions targeting compliance of vaccinated and non-vaccinated individuals. The original variant of the virus circulates. All panels show relative difference in the cumulative number of new infections as compared to the no-vaccination scenario. **a** and **b** Vaccination rollout not supplemented with compliance interventions three and six months into the vaccination rollout, respectively. **c** and **d** Vaccination rollout supplemented with compliance interventions targeting non-vaccinated individuals three and six months into the vaccination rollout, respectively. **e** and **f** Vaccination rollout supplemented with compliance interventions targeting vaccinated individuals three and six months into the vaccination rollout, respectively. **g** and **h** Vaccination rollout supplemented with compliance interventions targeting both vaccinated and non-vaccinated individuals three and six months into the vaccination rollout, respectively. Magenta curves mark boundaries between parameter regions with different sign of the cumulative number of new infections. The scale of x-axis is not linear since the axes were obtained by conversion of the vaccine uptake rate to the vaccination coverage following three and six months after the start of the vaccination rollout.

The vaccine efficacy where the vaccination campaign does not cause excess infections due to the reduction of compliance is smaller for more transmissible strains, in particular the Deltalike variant (Figures 5**a** and 5**b** and Figures **1a, 1b**, **2a**, and **2b** in Supplementary materials).

We refer to the analyses above as to the epidemic dynamics without compliance-targeted interventions. In the following section, we investigated the impact of interventions targeted at maintaining compliance with physical distancing; we compared this to the epidemic dynamics without compliance-targeted interventions and the scenario without either vaccination or interventions.

### Interventions targeting compliance

To investigate how interventions may improve the impact of vaccination rollout, we considered an intervention that targets compliance of those who are not yet vaccinated and an intervention targeted at the vaccinated population. We assume that the first intervention targets non-vaccinated individuals and is successful in keeping the duration of compliance at the pre-vaccination length (30 days) as vaccination coverage grows. The second intervention, targeted at vaccinated individuals, succeeds in convincing vaccinated individuals to abstain from increasing the contact rate above that of the contact rate of non-compliant individuals. Our model predicts that a successful implementation of either of these interventions reduces the cumulative number of new infections after vaccination rollout and can get this number below the level of the no-vaccination scenario. The effectiveness of these interventions depends on the circulating variant and the vaccine uptake rate. We summarize our findings in Figures 5 and Figures **1** and **2** in Supplementary materials.

#### Intervention 1: targeting compliance of nonvaccinated individuals

For all three variants, an intervention that targets compliance of non-vaccinated individuals (Figures 5**c** and 5**d** and Figures **1c, 1d**, **2c**, and **2d** in Supplementary materials), reduces the minimal efficacy of vaccine required for the cumulative number of new infections after three and six months following the vaccination rollout to be smaller than in the no-vaccination scenario at the respective time points. Moreover, the minimal vaccine efficacy after six months of the vaccination rollout is lower than the minimal efficacy after three months. Above the magenta curve, the reductions in the cumulative number of new infections are higher as compared to the scenario where vaccination rollout is not supplemented with compliance-targeted intervention (Figures 5**a**, 5**b**, 5**c**, and 5**d**). If there is an excess of new infections, it is smaller as compared to the no compliance targeted intervention scenario (Figures 5**a**, 5**b**, 5**c**, and 5**d**). Finally, we compare the performance of this intervention across different variants. While for all variants the intervention lowers the vaccine efficacy minimum at which the cumulative number of new infections decreases compared to the no-vaccination scenario, this threshold becomes lower for more transmissible variants. Similarly, the intervention yields larger relative reductions in the cumulative number of new infections for more transmissible variants.

#### Intervention 2: targeting compliance of vaccinated individuals

Effects of this intervention on the cumulative number of new infections depend on the circulating virus variant, vaccine efficacy, and vaccination rate (Figures 5**e** and 5**f**).

For the original variant and a slow vaccination rate, we observe that after three months of vaccination for the whole range of vaccine efficacies that were considered there is excess of infections as compared to the no-vaccination scenario (Figure 5**e**). This is contrary to the scenario when the vaccination rollout is not supplemented with compliance-targeted interventions, where vaccinated individuals are characterized by the increased contact rate. This outcome occurs due to the change in mixing. As vaccinated individuals have less contacts, more transmission contacts occur in the non-vaccinated population leading to the increase in the number of infections.

At the same time point but given a fast vaccination rate, we also see mixed results. For the combination of the vaccine efficacy and vaccination rate which gives a decrease in the cumulative number of the new infections in the scenario where vaccination rollout is not supplemented with compliance-targeted intervention, we see this decrease reducing in magnitude. On the other hand, the minimum of vaccine efficacy where the cumulative number of new infections is lower. Finally, the region with excess infections is smaller compared to the scenario where the vaccination rollout is not supplemented with compliance-targeted interventions.

Six months after start of the vaccination rollout, the situation is similar (Figure 5**f**). Given a slow vaccination rate, the minimum of vaccine efficacy where the relative increase of infections can be avoided is higher than in the scenario where the vaccination rollout is not supplemented with the intervention. But if the vaccination rate is fast, than the respective vaccine efficacy minimum is lower than it was without the intervention.

The dynamics for different regions of the vaccine efficacy and vaccination rate for an Alpha-like or a Delta-like variants when the intervention is deployed are qualitatively similar to the dynamics of the original strain (Figures **1e, 1f**, **2e**, and **2f** in Supplementary materials).

### Combination of two interventions

Finally, combination of the two compliance-targeted interventions leads to improvements that exceed the effects of individual interventions (Figures 5**g** and 5**h** and Figures **1g, 1h**, **2g**, and **2h** in Supplementary materials). For all three variants, the minimum for vaccine efficacy where the excess of infections as compared to the scenario without compliance-targeted intervention can be avoided, decreased. Also, excess in the cumulative number of infections decreased for the region of vaccine uptake rate and vaccine efficacy that we considered. Similar reductions relative to the scenario where the vaccination rollout is not supplemented with compliance-targeted intervention are achieved for the more transmissible variants, provided a fast vaccination rate (Figures **1g, 1h**, **2g**, and **2h** in Supplementary materials).

### Sensitivity analysis of the epidemic dynamics with vaccination

We performed sensitivity analysis to test the robustness of our findings to the variations of the initial conditions and parameter values. More specifically, we were interested whether the possibility of a relative excess in the cumulative number of new infections is preserved and how its value changes. In our analyses we varied one parameter at a time while keeping the initial conditions and parameter values fixed to the values that were used to produce simulations summarized on Figures 3 and 4. We considered the dynamics of the original variant. For the ranges used in Sensitivity analysis see Table 1.

The existence of the relative increase in the cumulative number of infections after three and six months of the vaccination rollout as compared to the no-vaccination scenario is preserved across the intervals for the initial conditions that we considered (Figures **6**-**9** in Supplementary materials). The size of the relative increase in the cumulative number of infections varies continuously, with the largest increase in its value as compared to what we observed in the main analysis (Figure 3) being equal approximately to 20%. The relative difference in the cumulative number of new infections has the highest sensitivity to the initial state of the seroprevalence as compared to the initial conditions in other compartments.

Increase in the relative difference in the cumulative number of new infections as compared to no vaccination scenario is preserved on the sampled intervals for the duration of the exposed and infectious period (Figure **11** in Supplementary materials). In the slow vaccination rate scenario, the absolute size of the cumulative number of new infections after three and six months after the start of the vaccination rollout is not sensitive to perturbations in either parameter (Figures **10a** and **10c** in Supplementary materials). On the other hand, the cumulative numbers of new infections in the fast vaccination scenario are very sensitive to the the duration of the infectious stage but not to the the duration of the exposed stage (Figures **10b** and **10d** in Supplementary materials).

Both the absolute value of the cumulative number of new infections and the relative difference in the number as compared to the no-vaccination scenario are very sensitive to the contact rates of compliant and non-compliant individuals (Figures **12** and **13** in Supplementary materials). The cumulative number of new infections after three months of the vaccination rollout is more sensitive to both than the cumulative number after six months. The cumulative number of new infections is increasing when either one of the contact rates is growing and is highest in the scenario where the average contact rate of the population was close to the pre-pandemic level. The cumulative number of new infections decreases below no-vaccination scenario when the contact rate of compliant individuals is sufficiently close to the contact rate of non-compliant individuals. This happens since the average contact rate of non-vaccinated individuals does not grow significantly even as the growing vaccination coverage causes a decrease in the proportion of compliant population. From Figure 4 we recall that when the original variant circulates, new infections occur mainly among non-vaccinated individuals. This is different for a more transmissible variant such as Delta, where vaccinated individuals contribute significantly to the cumulative number of new infections.

We observe that there is an excess of infections relative to the no-vaccination scenario for the whole range of values for the contact rate of non-compliant individuals that we considered (Figure **13** in Supplementary materials). The largest relative increase in the cumulative number of infections happens when the contact rate of non-compliant individuals is close to the prepandemic levels and the contact rate of compliant individuals is significantly lower. Therefore since the growing vaccination coverage causes modifications of the compliance distribution, the average contact rate in the non-vaccinated individuals increases significantly.

The cumulative number of new infections and the relative difference in the cumulative number of new infections (as compared to the no-vaccination scenario) are very sensitive to variations of the rate of moving to the compliant state and the duration of the compliant state (Figures **14** and **15** in Supplementary materials). The cumulative number of infections is the highest when individuals move to the compliant state at a slow rate but the duration of the compliant state is low (Figure **14**). As the rate of moving to compliant state and the duration of being compliant increase the, cumulative number of infections decreases. The relative difference in the cumulative number of infections has the opposite relationship with the two parameters (Figure **15**). Such that, the difference is largest when the rate of moving to compliant state is fast and the average duration of staying of compliant is long. We observe that the duration of compliant state has little effect on the possibility of excess infections as compared to the no-vaccination scenario. However, if the rate of moving to compliant state is sufficiently high, the cumulative number of infections will exceed the no-vaccination scenario level.

For the description of methodology and the complete treatment of this topic see Supplementary Materials.

### Supplementing vaccination rollout with a lockdown

Our simulations indicated that due to compliance waning as the vaccination coverage grows, it is possible that an additional prevalence peak appears. So far, in our simulations no centralized intervention triggered by a steep increase in the number of new cases was modeled. Here we consider such an intervention, whereupon if during the vaccination rollout the prevalence of new infectious cases exceeds a certain threshold, the government tightens the lockdown, further restricting the average contact rate. Once the prevalence falls bellow the threshold, the lockdown is being relaxed to its prior state. We investigated the effect of the threshold prevalence at which the lockdown is initiated on the cumulative number of new infections three and six months after the start of the vaccination campaign (Figures **3**-**5** in Supplementary materials).

Our simulations indicate that supplementing the vaccination rollout with lockdown which initiates once the prevalence of infectious cases exceeds a threshold can prevent increase of the cumulative number of new infections after three and six of the vaccination rollout as compared to no-vaccination scenario (Figure **3** in Supplementary materials). The cumulative number of new infections after three months of the vaccination rollout are larger for the fast vaccination rollout than for the slow. Interestingly, the cumulative number after three months of the vaccination rollout for either vaccination rate is not sensitive to changes in the lockdown strengthening/relaxation threshold on the range that we consider. On the other hand, the cumulative number of new infections after six months of the vaccination rollout for both slow and fast vaccination rates is increasing as the threshold for the strengthening/relaxation of the lockdown grows. Finally, we observe that the relative decrease in the cumulative number of infections is higher for the cumulative number of infections six months after the rollout than after three. While the relative difference in the cumulative number of new infections is larger for a lower lockdown initiation threshold, the gain is not sufficiently large to warrant a strict lockdown that initiates early. The largest decrease in the cumulative number of new infections relative to the no-vaccination scenario happens when the vaccination rate is fast and the vaccine efficacy is high. Decreasing in either one of these parameters causes the relative difference to decrease (Figure **4** in Supplementary materials). On the other hand, the largest decrease in the cumulative number of new infections relative to the vaccination rollout without compliance interventions happens when the vaccination rollout is fast and the vaccine efficacy is low (Figure **5** in Supplementary materials).

In summary, we gain the following insights for different vaccination strategies and virus variants: (a) if vaccinated and non-vaccinated individuals relax their compliance with physical distancing measures, the cumulative number of new infections may be higher than the no-vaccination scenario, regardless of the vaccine uptake rate; (b) Fast vaccine uptake rate may not always be advantageous. If the efficacy is very high, than fast vaccine uptake rate will lead to reduction of infections relative to the no-vaccination level. If the efficacy is low, fast vaccine uptake rate combined with diminished compliance may lead to a significant relative increase in the number of infections; (c) For all variants that we considered, an intervention targeting the non-vaccinated population is effective in reducing the number of infections below the no compliance-targeted intervention scenario and reduces the minimum value for vaccine efficacy necessary to lower this number below the no-vaccination scenario level; (d) The intervention that targets compliance of non-vaccinated individuals yields better results in a long run than in short run, with the threshold vaccine uptake rate at six months significantly lower than it was at three months; (e) Slow vaccination with a combined compliance-targeting intervention can reduce numbers of infections as compared to the no-interventions scenario. But in order to reduce the number of infections below the level of the no-vaccination scenario, vaccine efficacy should exceed 65%; fast vaccination with a combined intervention reduces the number of new infections even for lower vaccine efficacy.; (f) Strengthening of the lockdown triggered by the rise in prevalence is another intervention that can prevent increase in the cumulative number of new infections. Our results indicate that the initiation threshold for the lockdown can be sufficiently high, thus potentially allowing for shorter periods of the slowing down of the economy.

## Discussion

Using a compartmental model for the spread of SARS-CoV-2 in a population, where physical distancing measures are in place, we investigated the impact of declining compliance with physical distancing measures as vaccination is rolled out on the numbers of infections. One of the key features of our model is a distinct treatment of the loss of compliance by vaccinated and non-vaccinated populations, each of which can relax the compliance of physical distancing measures to a different degree. Additionally, we extended the compliance process to the whole population and not only the susceptible individuals, which qualitatively affects mixing patterns in the population.

Our main finding is that, if compliance decays as the vaccination coverage grows, the speed of vaccination rollout has a strong impact on whether the cumulative number of new infections can be decreased three and six months after the start of vaccination below the level that would have been expected without vaccination. If vaccination rollout is slow, its positive effects on the incidence will be counteracted by fading compliance and increasing contact rates in the population. This may lead to an increase in the prevalence exceeding the prevalence in a situation without vaccination and, in the short term, we may even see an additional epidemic peak. If vaccination is rolled out faster, these detrimental effects can be avoided. The outcome will depend on the vaccine efficacy. If the efficacy is high, then the cumulative number of new infections will decrease relative to the no-vaccination scenario. If the vaccine efficacy is low and the vaccination rate if fast, an excess of infections is possible in the first six months of the vaccination rollout. Generally, given a low vaccine efficacy, our model predicts that after the first six months of the vaccination rollout, the cumulative number of new infections is higher for a faster vaccination uptake rate. This effect happens due to the loss of compliance by vaccinated individuals. Note that, since among the excess infections a certain proportion of infected people will have been vaccinated, they will have a low probability of developing severe disease or death. Finally, as a result of our comprehensive analysis of the effect of the vaccination rate and vaccine efficacy on the cumulative number of new infections, we derived threshold curves which separate parametric regions where the relative difference in the cumulative number of infections as compared to the no-vaccination scenario changes sign. We observed, that if the vaccine has a high efficacy, then the excess of infections can be avoided for a relatively low vaccination uptake rate. As the vaccine efficacy decreases, the uptake rate increases.

In their recently published work Gozzi et al (52) also considered the impact of the feedback between the epidemic dynamics, the vaccination rollout, and compliance with physical distancing on infection transmission dynamics. The authors investigated the effects of waning compliance due to the growing vaccination coverage provided different vaccination strategies and vaccine efficacies across populations with different age contact matrices. Both ours and Gozzi et al (53) qualitative findings are in agreement and are consistent with the results of the earlier studies that have shown that factors that contribute to drastic increase of contact rates (such as vaccination-related behavioral change or premature reduction/removal of nonpharmaceutical interventions) may reduce the benefits of a vaccination programme (54–57).

Motivated by the conclusions drawn by these studies, we considered the effect of supplementing the vaccination campaigns with communication strategies promoting maintenance of physical distancing behavior aimed at both vaccinated and non-vaccinated individuals and learned that 1) those interventions can significantly improve the outcome of vaccination campaign; 2) the choice of a specific information intervention should be informed by the epidemic circumstance of the situation (such as the dominant variant and speed of vaccination rollout).

An intervention that succeeds in maintaining the compliance with physical distancing in people not yet vaccinated on the same level as before the start of vaccination ensures significant decrease of the cumulative number of new infections throughout. Moreover, for all three virus variants, supplementing vaccination rollout with this intervention reduces the vaccine efficacy threshold for which the cumulative number of new infections is lower than without the vaccination. This effect is seen in both short and long term, but is more pronounced in the long term. The effect for an intervention that targets vaccinated individuals to prevent them from increasing their contact rates after being vaccinated depends on the transmissibility of the dominant variant. If the original variant circulates, the intervention has a positive impact for a fast rollout of vaccination, but cannot avoid detrimental effects of waning of compliance if the vaccination rollout is slow. On the other hand, if the dominating variant has the same transmissibility as Alpha or Delta, then the intervention can improve the outcome of the vaccination rollout over the no-vaccination scenario even when the vaccination rate is slow. Interestingly, for the original and an Alpha-like variant, given a slow vaccination rate, the minimum vaccine efficacy threshold required to avoid a surplus of infections is higher when the vaccination rollout is supplemented with the intervention than when it is not. If a Delta-like variant circulates, supplementing the vaccination rollout with the intervention reduces the threshold for all vaccination rates that we considered. Only the combined effect of both interventions can consistently reduce the cumulative number of new infections below the level of the no-vaccination scenario regardless of the rollout speed (in the vaccination rate range that we considered).

Finally, we compared the effect of compliance-targeting interventions with a centralised intervention that mimics tightening/relaxation of the lockdown when a prevalence threshold is crossed. We observed that the possibility of an excess of infections is eliminated and yields larger decreases in the cumulative number of new infections over the no-vaccination scenario than the compliance-targeting interventions, both in the short term and in the long term. The outcomes of supplementing the rollout with this intervention are not sensitive to the prevalence threshold. However, it may come at a price of disrupted social fabric and slowing down of the economy.

Our results are based on some simplifying assumptions, one of them that physical distancing measures remain in place throughout the time period of analysis (six months). While this would be advantageous for preventing transmission of the virus, it might not be feasible out of societal and economic reasons. Therefore, compliance rates may wane even faster in real populations and contact rates may be up to higher, possibly pre-pandemic values during the rollout of vaccination. We do not expect that this would change our results much, as our results are obtained relative to the no-vaccination scenario, which would similarly be affected by a change in physical distancing measures. We expect therefore that the relative effects of vaccination would remain similar as in our simulations. We also assumed that the speed of vaccination rollout stays constant over the time period of six months, which is not the case in reality. In the Netherlands for example, vaccination rates have increased substantially after a slow start in January 2021 (38). These rates will depend on many factors, nevertheless large differences will remain between countries. Finally, we have captured the dependence of rates of becoming compliant and non-compliant on the incidence of new infectious cases and vaccination coverage, respectively, using linear functions. As the vaccination in many countries continues and the population response data is collected, a more precise formulation of the response functions can be obtained. However, our results predominantly depend on the assumed monotonicity of these functions.

Furthermore, our model is relatively simple, not taking into account age structure and heterogeneity in contact patterns. Therefore, we do not attempt to make quantitative predictions on the impact of vaccination, but we provide qualitative insight into possible effects of waning compliance with physical distancing in the face of increasing vaccination coverage.

A number of studies/reports estimated the bounds for vaccine efficacy for the original variant in terms of reducing the infection for some vaccines approved for use in Europe (21, 22, 24, 25). As Alpha (B.1.1.7) and Delta (B.1.617.2) variants emerged and, in turn, became dominant in many European countries, the first estimates for vaccine efficacy for reducing the infection became available (30, 31, 58). Whether the reduction in infection comes in the guise of reduction of susceptibility or transmissibility of vaccinated individuals is not known. Therefore, in this work we modeled the vaccination to be all-or-nothing and vaccine efficacy was given in terms of probability of conferring full protection from becoming infected. Our sensitivity analyses (Figure 5 and Figures **1**, **2**, **4**, and **5** in Supplementary materials) show that the effect of a vaccination campaign and of individual interventions is highly sensitive with respect to this parameter. However, we observed that if no compliance-targeting interventions accompany the vaccination rollout, the range of efficacies for which a surplus of new infections as compared to no-vaccination is possible three and six months following the vaccination rollout falls within the vaccine efficacy boundaries that were reported for different vaccines (22, 24–27, 29–31, 59). To implement the most efficient vaccination rollout it is important to know the boundaries of vaccine-conferred reduction of transmission.

Finally, in this work we have considered dynamics of circulation of three SARS-CoV-2 virus variants, the original variant and two mutations, whose transmission potential is similar to the Alpha and Delta variants. For all three variants, we modeled the immunity induced by the vaccine to be of the identical type (sterilising).

Our results also show that speed of rollout of a vaccination campaign is important, because the speed of the rollout and subsequent changes in contact rates strongly impact cumulative number of new infections. Although in the scenario where vaccination rollout is fast the population may fair worse than it would have been without vaccination in the short term especially for a more transmissible virus variant -on the longer term (*>* 1 year) it has vast advantages in terms of numbers of infections prevented.

Our results emphasize the importance of communication by public health professionals on continued adherence to self-imposed measures, to those who are awaiting vaccination as well as to those already vaccinated. Communication messages need to be different and targeted specifically to these two groups. We highlight the positive overall effects of vaccination campaigns in combination with continued adherence to non-pharmaceutical preventive measures.

### Model

We developed a compartmental deterministic model that describes SARS-CoV-2 transmission and vaccination roll-out in a population. Subsequently, we modified this model to include acquisition and loss of compliance with physical distancing measures as individuals continuously get exposed to information about disease spread as well as the progress of vaccination rollout (Figure -1). We informed the model using parameter values from the literature as well as estimating parameters from publicly available data for the Netherlands, Belarus, Denmark, and Israel. We used the model to investigate the effects of interactions between disease transmission, vaccination rollout, and changing compliance with physical distancing measures on transmission dynamics.

#### Population compartments

The SARS-CoV-2 transmission dynamics follow a Susceptible-Exposed-Infectious-Recovered (SEIR) framework that divides the population into the following compartments: susceptible (*S*), latently infected (also referred to as “exposed”, *E*), infectious (*I*), and recovered (*R*). Susceptible individuals (*S*) become latently infected (*E*) with rate *λ*_inf_ proportional to the fraction of infectious individuals (*I/N*, where *N* is the total population size). Individuals stay latently infected (*E*) for an average duration of 1*/α* days after which they become infectious (*I*). Infectious individuals recover after 1*/γ* days and move to compartment *R*. Because of a relatively short time horizon of our analyses (not exceeding six months) and relatively small case fatality ratio, we disregarded demographic processes such as births and deaths, and therefore the population size *N* is constant. Additionally, we assumed that once individuals recover they acquire permanent immunity and cannot be re-infected. Since we are interested in understanding the qualitative dynamics that follow from interaction of infection transmission, changes in compliance, and vaccination rollout, we did not consider different outcomes of infection with SARS-CoV-2 (e.g., asymptomatic or symptomatic infection, hospitalisation, death etc.). The infectious compartment (*I*), therefore, contains individuals who are asymptomatic, or have mild or severe symptoms.

The dynamics of infection transmission are modelled for three variants of the SARS-CoV-2 virus: first, the original variant that was predominant in Europe prior to fall 2020; second, the more transmissible Alpha (B.1.1.7) variant, that was initially detected in the UK and became dominant in many European countries in the spring of 2021; and finally, the “hyper-contagious” Delta (B.1.617.2) variant, which became dominant in Europe in summer 2021. We parameterized the differences between these variants by using different probabilities of transmission per contact, *E*. We assumed that in all other respects the variants have the same properties. We investigated model dynamics where only one of the three variants circulates in the population.

To model vaccination, the population was stratified into vaccinated and non-vaccinated classes. While for some vaccines authorised for use in Europe (BioNTech/Pfizer, Moderna and AstraZeneca, (17)), two vaccine doses, as well as a certain time period passing after the second dose are required for full immunisation, we modelled vaccination as a single event that confers protection instantaneously. We assumed that individuals do not obtain a diagnostic or antibody test prior to vaccination, and therefore infected and recovered individuals also get vaccinated. Thus, individuals in all epidemiological compartments can get vaccinated, but only those who were susceptible (*S*) at the time of vaccination may become immunised (*V*). The vaccination rate of susceptible, exposed, and recovered individuals is denoted by *υ*. We introduced a parameter *k*_1_, 0 *≤ k*_1_ *≤* 1, such that *k*_1_*υ* denotes the vaccination rate for individuals in the infectious compartment, to reflect that a fraction of infectious individuals (who have symptoms) might not be eligible for or might decide against vaccination. In the main analysis we considered the case where infectious individuals get vaccinated at the same rate as individuals in other compartments (*k*_1_ = 1). We explored sensitivity of the dynamics to variation of *k*_1_ and observed little effect of changes in this parameter (the Supplementary materials). We assumed that the vaccine works as all-or-nothing, i.e. upon vaccination, a proportion *ω* of susceptible individuals (*S*) is fully protected (*V*), while in a proportion 1 *− ω* of susceptible individuals the vaccine has no effect. We refer to *ω* as “vaccine efficacy” in the context of conferring sterilising immunity. Vaccination does not confer protection to individuals, who were in other infection compartments (*E, I* and *R*) at the time of vaccination, and their infection progression is identical to that of non-vaccinated individuals. Individuals who were vaccinated but did not obtain the protection are denoted by *S*^*V*^, *E*^*V*^, *I*^*V*^ and *R*^*V*^.

Studies based on data collected in Israel estimated that the BNT162b2 mRNA COVID-19 vaccine developed by Pfizer/BioNTech reduced the acquisition rate for asymptomatic SARS-CoV-2 infection by 80% (26) up to 95% (27). Similarly high efficacy against infection acquisition were reported for the mRNA-1273 COVID-19 vaccine developed by Moderna, NIAID (28). For the adenovirus Ad26.COV2.S COVID-19 vaccine developed by Janssen Pharmaceutical Companies the efficacy in preventing infection with SARS-CoV-2 is reported to be 76% (29). These results were estimated from data collected between December 2020 and April 2021 in the USA. During this period, the original variant and the Alpha variant (B.1.1.7) were the dominant circulating variants. Since then, the “hyper-contagious” Delta variant (B.1.617.2) has become dominant in many European countries (10) and the USA (11). A recent study based on data from Israel estimated a significant reduction of BNT162b2 efficacy for the Delta variant in preventing infection, which was 64% after two doses (30). This estimate was supported by another report based on the data in a highly vaccinated health system workforce of California San Diego Health (31). Therefore, in our analyses, we varied *ω* in the range of 0.4 and 1.0.

Finally, in addition to infection status and vaccination status, individuals in the model are either compliant or non-compliant with physical distancing measures (compliant compartments denoted by superscript *C*: *S*^*C*^, *E*^*C*^, *I*^*C*^, and *R*^*C*^). Compliant individuals thus have on average a lower contact rate than non-compliant individuals; both contact rates are assumed to be lower than pre-pandemic levels. We denote the contact rate of non-compliant individuals by *c*, and define a reduction factor *r*_1_ that describes the reduction in contact rate of compliant individuals compared to non-compliant individuals, so 0 *≤ r*_1_ *≤* 1. Transitions between the compliant and non-compiant state are described by a modelling framework similar to Perra et al (41) and previously used in (43). We modeled the compliance acquisition rate, *λ*_*C*_, as a function of the incidence of infection, assuming that individuals obtain information about numbers of cases through mass-media and health authorities. We assumed that compliance wanes when case numbers drop or when the disease is no longer present, and individuals return to the non-compliant state at rate *µ*. If there is no vaccination programme in place then this rate, *µ*, is constant. However, if vaccination rollout is in progress and as vaccination coverage increases, individuals may feel less motivated to comply with physical distancing measures; we implemented this effect by taking *µ* as a linear function of vaccination coverage, i.e. the rate of losing compliance increases with increasing vaccination coverage. We assumed that only non-vaccinated individuals can be in the compliant state, while vaccinated individuals move into a separate non-compliant state permanently, and even have higher contact rates than non-vaccinated non-compliant individuals. We use *r*_2_ *≥* 1 to denote the increase in the contact rates of vaccinated individuals relative to the contact rate of non-compliant individuals, *c*. Compliant individuals get vaccinated at the same rate as non-compliant individuals. All individuals who were vaccinated will have the same (increased) contact rate regardless of whether vaccination was successful.

#### Rates

In this section we define the transition rates that depend on the incidence of infectious cases and on vaccination coverage: rates of infection acquisition, and rates of acquisition and loss of compliance.

We assumed that individuals become infected at a rate that depends on the fractions of different types of infectious individuals, as well as on the mixing of compliant, non-compliant and vaccinated individuals. Therefore, infection acquisition rates as well as infection transmission rates depend on compliance and vaccination status of susceptible and infectious individuals. We define the following matrix to summarize transmission rates between different types of susceptible and infectious individuals.

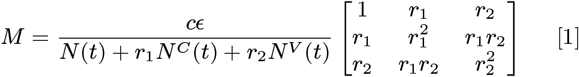

with

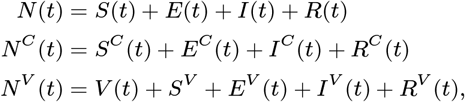

where [*M*]_11_ captures the transmission of infection from non-compliant *I* to non-compliant *S*, [*M*]_12_ from compliant *I* to non-compliant *S*, and [*M*]_13_ from vaccinated *I* to non-compliant *S*. Similarly, the second row of the matrix captures the transmission of infection to susceptible individuals who are compliant, *S*^*C*^. Finally, the third row of the matrix captures the transmission of infection to individuals who are susceptible despite vaccination, *S*^*V*^.

We assumed that as individuals learn about new infections they become compliant with physical distancing measures, and therefore compliance is gained at a rate *λ*_*C*_ which is a positive increasing function of the incidence of infectious cases (equal to the rate with which individuals leave the exposed stage):

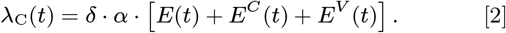

We assumed that compliance is not permanent, becoming shorter as the vaccination coverage grows, and thus we model compliant state to have an average duration 1*/µ*, such that *µ* is a positive increasing function of the vaccination coverage, 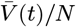 :

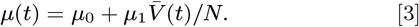

#### Equations

The system of ordinary differential equations (4) provides a full description of the model.

Dynamics of non-compliant individuals:

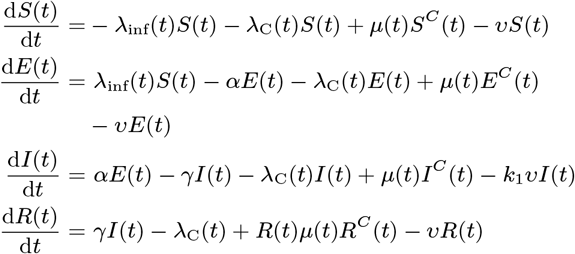

Dynamics of compliant individuals

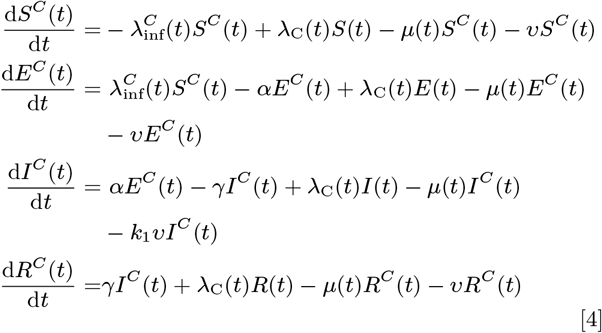

Dynamics of vaccinated individuals:

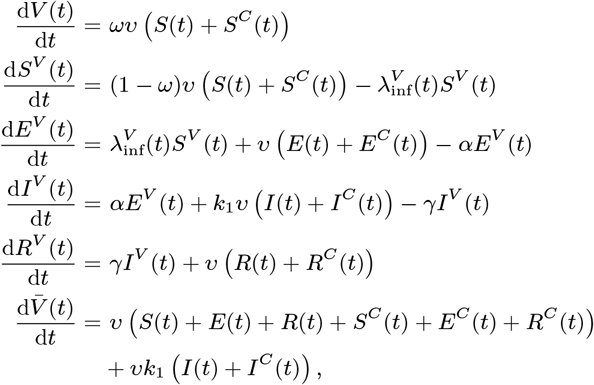

where

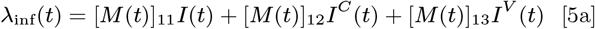

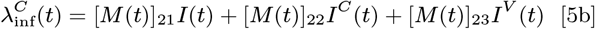

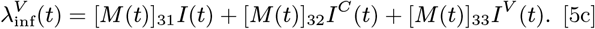

### Parameters and initial data

A full list of parameters and their values is given in Table 1. Here we elaborate on our choice of initial conditions, as well as on the chosen values of the behavioral parameters.

#### Initial data

To model the dynamics of SARS-CoV-2 we used the approximation made by RIVM for the week November 11-17 for the number of infectious individuals and set the total number of currently infectious individuals, *I* + *I*^*C*^, at the start of vaccination rollout to 112,435 (4). We have used this value in the main analysis and performed sensitivity analysis to investigate the sensitivity of our results to this choice. To estimate the fraction of recovered individuals, *R* + *R*_*C*_ we used seroprevalence data from a serological study conducted in the Netherlands in June and July of 2020. In this study, based on an age-stratified and regionally weighted representative sample of the Dutch population, the seroprevalence was estimated at 4%. In a later update of that study, seroprevalence was found to be 14% in February 2021 (46, 47). We set the number of recovered, *R* + *R*^*C*^ such that at the start of vaccination rollout, which was in January 2021 in the Netherlands, the fraction of recovered in the population was 8%, and performed sensitivity analysis with respect to this initial value.

According to the parameter values that we have selected the combined average duration of latent and infectious stage is estimated to be 11 days (Table 1, (60, 61, 63, 65)).

To estimate the total number of exposed individuals *E* + *E*^*C*^ at the start of the vaccination rollout, we assumed that, at the time, the epidemiological dynamics are in (pseudo) equilibrium, with the prevalence of infectious cases equal to 112,435 individuals (4). Using the average duration of infectious period equal to 7 days (64), we estimated that, at the start of the vaccination rollout, the daily incidence of new cases was 16,062 individuals. Using the average duration of the exposed period of infection equal to 4 days (60, 61, 63), we obtained *E* + *E*^*C*^. Having fixed the size of susceptible (*S* + *S*^*C*^), exposed (*E* + *E*^*C*^), and recovered (*R* + *R*^*C*^) compartments and using the total population size of the Netherlands, the size of the susceptible compartment (*S* + *S*^*C*^) follows.

We have set the initial proportion of compliant individuals to 65%. This was based on data on the compliance with maintaining a distance of 1.5m, from a study on behavioral measures and well-being conducted between November 11-15, 2020 (48) in the Netherlands.

We obtain

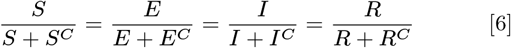

Using Eq. (6) and the percentage of compliant population, initial values for *S, E, I, R, S*^*C*^, *E*^*C*^, *I*^*C*^, *R*^*C*^ follow.

Setting the total population size to be equal to approximately that of the Netherlands, 1.7 *×* 10^7^ we obtain the initial data:

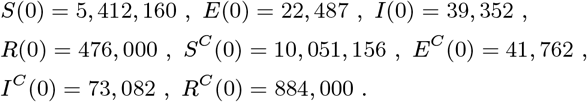

The initial values for the remaining compartments are set to 0.

#### Contact rates

We defined a contact as an encounter with another individual that is sufficiently long to have a conversation, or that involves physical interactions (51). The pre-pandemic contact rate in the Netherlands was reported to be equal to 14.9 individuals per day (51). We assume that the population is in the state of a partial lockdown at the start and throughout the vaccination rollout. In addition to the lockdown-related changes in the contact rate, individuals may reduce the contact rate further by complying with government-recommended physical distancing measures (e.g. work from home as much as possible). A fraction of the population is more compliant with these physical distancing measures and the remaining fraction is less compliant, such that contact rates in the compliant and non-compliant states are constant and the average contact rate is lower than pre-pandemic contact rate. However, as a consequence of vaccination and subsequent loss of compliance the average contact rate in the total population will change in time.

We fixed the contact rates for compliant and non-compliant individuals such that the effective reproduction number *R*_*e*_ at the start of the vaccination rollout was 1.1, which is in agreement with the estimate of *R*_*t*_ reported for the Netherlands in November 2020 (49). We calculated *R*_*e*_(0) assuming that *R*_0_ =*β/γ*= *ĉϵ/γ*=2. 5(60, 61).

Recall that the contact rates of non-compliant and compliant individuals are denoted by *c* and *r*_1_*c*. We calculated the effective reproduction number using the method described in (62) as

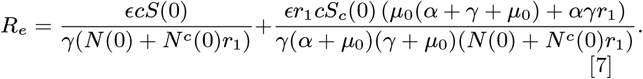

The value *R*_*e*_ = 1.1 is obtained for pairs of contact rates of non-compliant individuals, *c*, and compliant individuals, *r*_1_*c* (Figure 7).

**Fig. 7.**
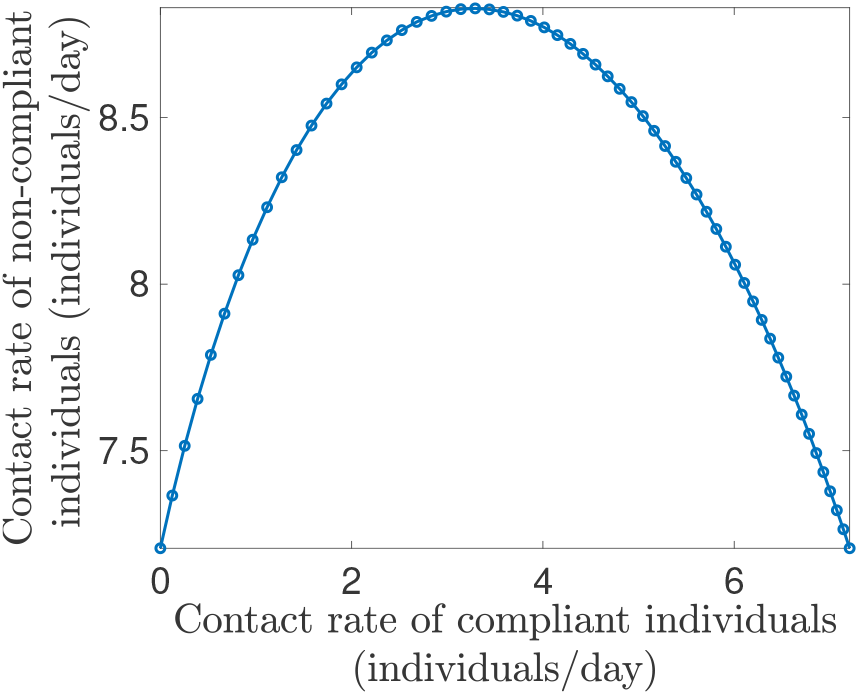
Pairs of contact rates of non-compliant and compliant individuals *c* and *r*_1_*c* such that effective reproduction number is equal to 1.1.

Of all pairs of contact rates that satisfy *R*_*e*_(0) = 1.1, we selected a combination such that the weighted average contact rate for the population at the start of the vaccination is 5 contacts per day. This value exceeds the reported number of contacts in the Netherlands during the government-imposed physical distancing measures in March 2020 by 1.5 contacts but is lower than the reported contact rate of 8.8 per day that was observed in June 2020, when some of the physical distancing measures were relaxed (51). The chosen parameter pair is *c* = 8.8 and *r*_1_*c* = 2.8.

Contact rates of vaccinated individuals were taken to be 1.5 times the contact rate of non-compliant individuals, assuming that after vaccination individuals will nearly return to the pre-pandemic contact behaviour.

#### Compliance

The proportion of compliant and non-compliant individuals in the population is determined by the compliance acquisition rate *δ* and compliance loss rate *µ*. For the main analysis we fixed the duration of compliance when there is no vaccination, 1*/µ*_0_ to 30 days. We selected the per capita rate of moving to the compliant state, *δ* = 4 *×* 10^*−*5^ so that given a constant daily incidence of 16,062 cases, 95% of the population is expected to be compliant. In the regime where the epidemic is seeded with the original variant in a population without any physical measures as much as 84% of the population can be compliant provided there were no compliant individuals at the start of the epidemic. This value denotes the case with high compliance acquisition rate. We investigated the sensitivity of the outputs to variation in per capita rate of moving to the compliant state and the compliance loss rate (Supplementary materials).

In the main analysis we considered a compliance decay scenario where as the vaccination coverage grows, the duration of compliance decreases, in particular when 33% of the population is vaccinated the compliant state lasts on average 7 days. In other words, for a daily incidence of 16,062 cases, which we used to initialize the model, and a slow vaccination rate, approximately 83% of the population is still compliant 6 months after start of the vaccination rollout, while for a fast vaccination rate only 32% are compliant (Figure 2**b**). These dynamics occur when the growth rate of compliant decay rate as the vaccination coverage increase is *µ*_1_ = 0.3 per day.

#### Model code

The model was implemented in MATLAB R2020b (66). The code producing the analyses and figures for this study is available at https://github.com/aiteslya/COVID-19-Vaccine-Compliance (67).

## Data Availability

All data and code referred to in the manuscript is deposited in the publicly available repository on GitHub.

https://github.com/aiteslya/COVID-19-Vaccine-Compliance

## ACKNOWLEDGMENTS

We thank Marc Bonten (UMC Utrecht) for comments on an earlier version of the manuscript. MEK acknowledges support from the Netherlands Organization for Health Research and Development (ZonMw; Grant no. 91216062 and Grant no. 10430022010001). GR acknowledges support from the Portuguese Foundation for Science and Technology (FCT; Grant no. 131_596787873). AT and HN acknowledge support from the Netherlands Organization for Health Research and Development (ZonMw; Grant no. 91216062).

