## Supplementary materials for "The importance of sustained compliance with physical distancing during COVID-19 vaccination rollout"

### 1 Compliance interventions for Alpha- and Delta-like variants

We investigated the sensitivity of the cumulative number of new infections to the vaccination uptake rate and vaccine efficacy in scenarios where the dominant SARS-CoV-2 virus variant is more transmissible than the original variant, i.e. when either an Alpha-like or a Delta-like variant circulates (Figures 1**a**, 1**b**, 2**a**, and 2**b**). For both variants, we also investigated the effects of interventions targeting compliance with physical distancing measures of vaccinated and non-vaccinated individuals (Figures 1**c**-1**h**, 2**c**- 2**h**)).

For both strains, the qualitative dynamics observed when vaccination rollout is not accompanied by additional interventions is similar to that of the original strain (Figures 1**a**, 1**b**, 2**a**, and 2**b**). More specifically, there is a region for vaccine efficacy and vaccination uptake rate, where the cumulative number of infections exceeds the number for the no-vaccination scenario three and six months after the start of the vaccination rollout. The highest increase above the numbers seen for the no-vaccination scenario is expected for a high uptake rate and low vaccine efficacy. Generally speaking, if the vaccination campaign is not accompanied by compliance-targeting interventions, to achieve a better result than the no-vaccination scenario, the vaccine efficacy should exceed a certain threshold. This threshold decreases with increasing vaccination uptake rate.

Similar to the original variant, the threshold vaccine efficacy is lower six months after the start of the vaccination rollout than it is after three. However, for the more infectious strains, the difference in the threshold vaccine efficacy is smaller than it was for the less infectious original variant. Finally, for both variants, in the regions where the cumulative number of infections exceeds that of the no-vaccination scenario, this excess is larger than it was for the original strain scenario (Figures **5a** and **5b** in the main text, Figures 1**a**, 1**b**, 2**a**, 2**b**).

For both variants, the intervention that targets compliance of non-vaccinated individuals, lowers the threshold vaccine efficacy as compared to the vaccination rollout without compliance-targeting interventions (Figures 1**c**, 1**d**, 2**c**, 2**d**). Similar to the threshold for the original variant, this curve is lower at six months than at three months.

The intervention targeting compliance of vaccinated individuals lowers the threshold vaccine efficacy as compared to the vaccination rollout without such intervention (Figures **5e** and **5f** in the main text, Figures 1**e**, 1**f**, 2**e**, 2**f**). For both Alpha-like and Delta-like variants, six months after start of vaccination, the threshold vaccine efficacy required to obtain improvements on the no-vaccination scenario has a more pronounced relationship with the vaccination uptake rate than it does after three months. Similar to the scenario when the original variant circulates, the threshold vaccine efficacy with low vaccination uptake rate is higher than when the vaccination rollout is not supplemented by compliance targeting interventions interventions.

Finally, the combination of the two interventions, yields the best results for either variant. However, the threshold vaccine efficacy is higher than for the original less-infectious variant (Figures **5g** and **5h** in the main text, Figures 1**g**, 1**h**, 2**g**, 2**h**).

### 2 Additional physical distancing intervention during the vaccination rollout

We considered a scenario where if during the vaccination rollout the prevalence of new infectious cases exceeds a certain threshold, the lockdown that we assumed was in place during the vaccination rollout becomes stricter, further diminishing the average contact rate. Once the prevalence falls bellow the threshold, the lockdown is being relaxed to its prior state. We refer to this intervention "dynamic" lockdown. We investigated the sensitivity of the outputs to the threshold prevalence at which the lockdown is initiated. The original variant of the virus circulates. The model parameters and initial conditions were fixed to the values used in the main text.

To perform the simulations we fixed the initial conditions and parameters to the values used in the main analyses. We assume that the lockdown reduces the average contact rate from 5 to 3 individuals per day. This is comparable to the number of contacts (3.5) residents of the Netherlands reported during the first weeks of the lockdown in March 2020 reported by Backer et al [1]. We considered the threshold for the initiation (and the relaxation) of the lockdown on the range of 50-1000 people. To assess the outcome of supplementing of the vaccination rollout with strengthening of the lockdowns we considered the following outputs: the cumulative number of new infections and the relative difference of the cumulative number of new infections as compared to the no-vaccination scenario where the lockdown is strengthened and relaxed in the similar way. The summary of our simulations are presented in Figure 3.

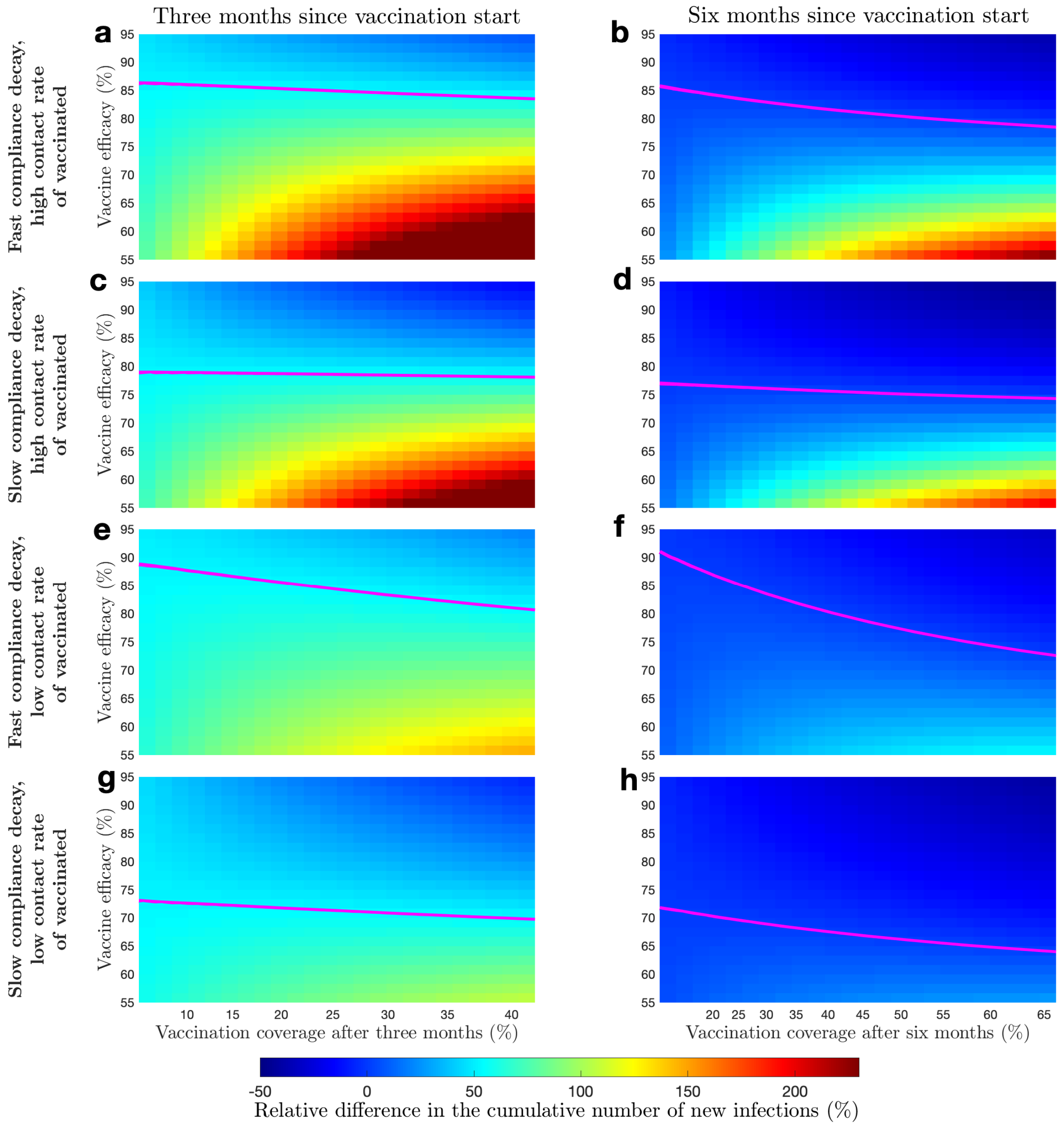

Figure 1: **Epidemic dynamics with and without interventions targeting compliance of vaccinated and non-vaccinated individuals.** An Alpha-like variant of the virus circulates. All panels show relative difference in cumulative number of new infections as compared to the no-vaccination scenario. **a** and **b** Vaccination rollout not supplemented with compliance interventions three and six months into the vaccination rollout, respectively. **c** and **d** Vaccination rollout supplemented with compliance interventions targeting non-vaccinated individuals three and six months into the vaccination rollout, respectively. **e** and **f** Vaccination rollout supplemented with compliance interventions targeting vaccinated individuals three and six months into the vaccination rollout, respectively. **g** and **h** Vaccination rollout supplemented with compliance interventions targeting both vaccinated and non-vaccinated individuals three and six months into the vaccination rollout, respectively. Magenta curves mark boundaries between parameter regions with different sign of the cumulative number of new infections. The scale of x-axes is not linear since vaccination coverage depends non-linearly on the vaccine uptake rate.

Both the cumulative number of new infections and the relative difference of the cumulative number as compared to the no-vaccination scenario is sensitive to the lockdown threshold value after six months of the vaccination rollout. In contrast, at three months after the vaccination rollout the threshold does not affects outcomes. After six months

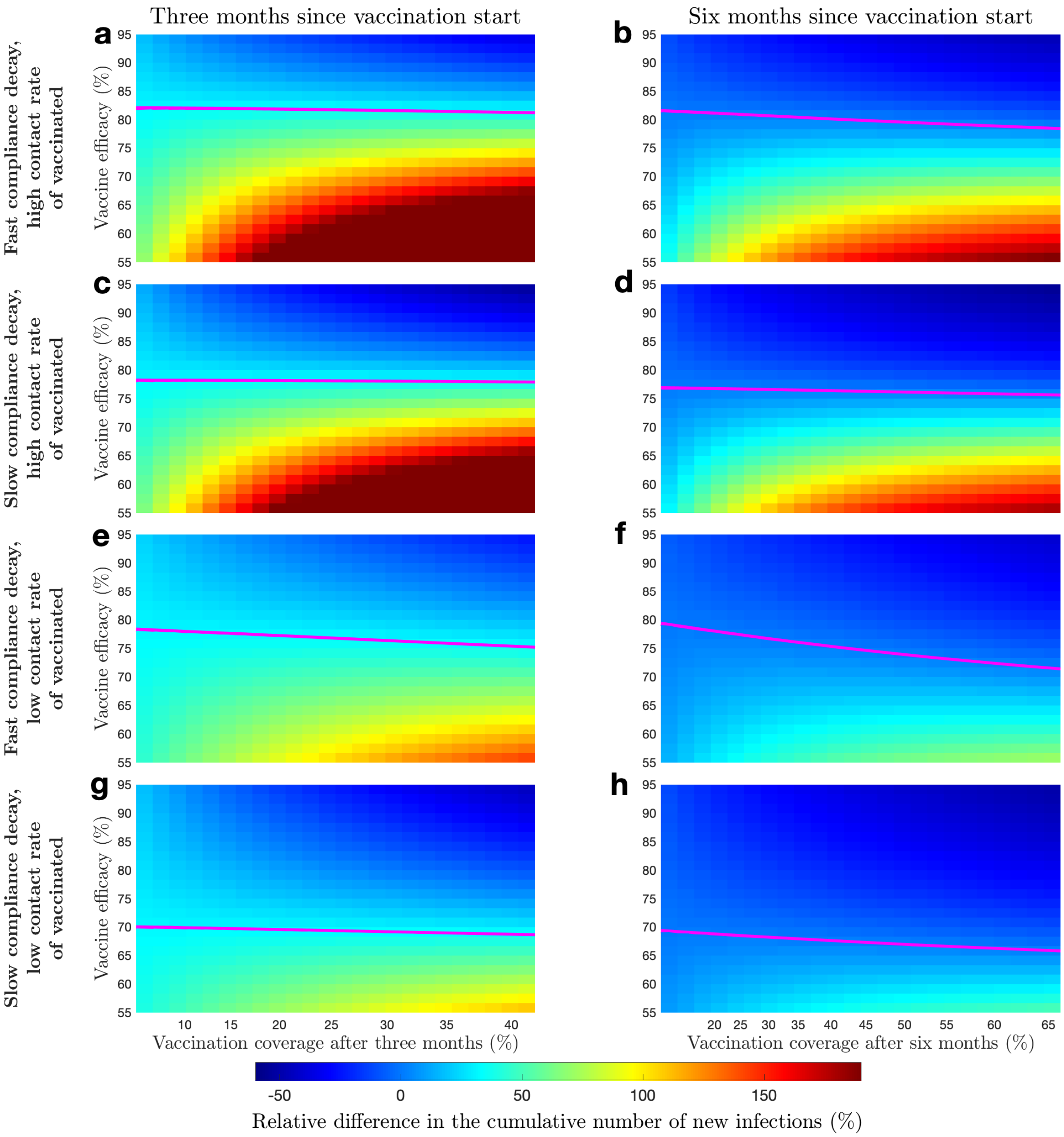

Figure 2: **Epidemic dynamics with and without interventions targeting compliance of vaccinated and non-vaccinated individuals.** A Delta-like variant of the virus circulates. All panels show relative difference in cumulative number of new infections as compared to the no-vaccination scenario. **a** and **b** Vaccination rollout not supplemented with compliance interventions three and six months into the vaccination rollout, respectively. **c** and **d** Vaccination rollout supplemented with compliance interventions targeting non-vaccinated individuals three and six months into the vaccination rollout, respectively. **e** and **f** Vaccination rollout supplemented with compliance interventions targeting vaccinated individuals three and six months into the vaccination rollout, respectively. **g** and **h** Vaccination rollout supplemented with compliance interventions targeting both vaccinated and non-vaccinated individuals three and six months into the vaccination rollout, respectively. Magenta curves mark boundaries between parameter regions with different sign of the cumulative number of new infections. The scale of x-axes is not linear since vaccination coverage depends non-linearly on the vaccine uptake rate.

of the vaccination rollout, we observe that as the threshold for initiation (and relaxation) of the lockdown increases, the cumulative number of new infections increases as well. However, when the vaccination rollout is supplemented with "dynamic" lockdown, the cumulative number of new infections is expected to decrease below the level of no-vaccination. It will decrease more for a fast vaccination rate than for a slow vaccination rate.

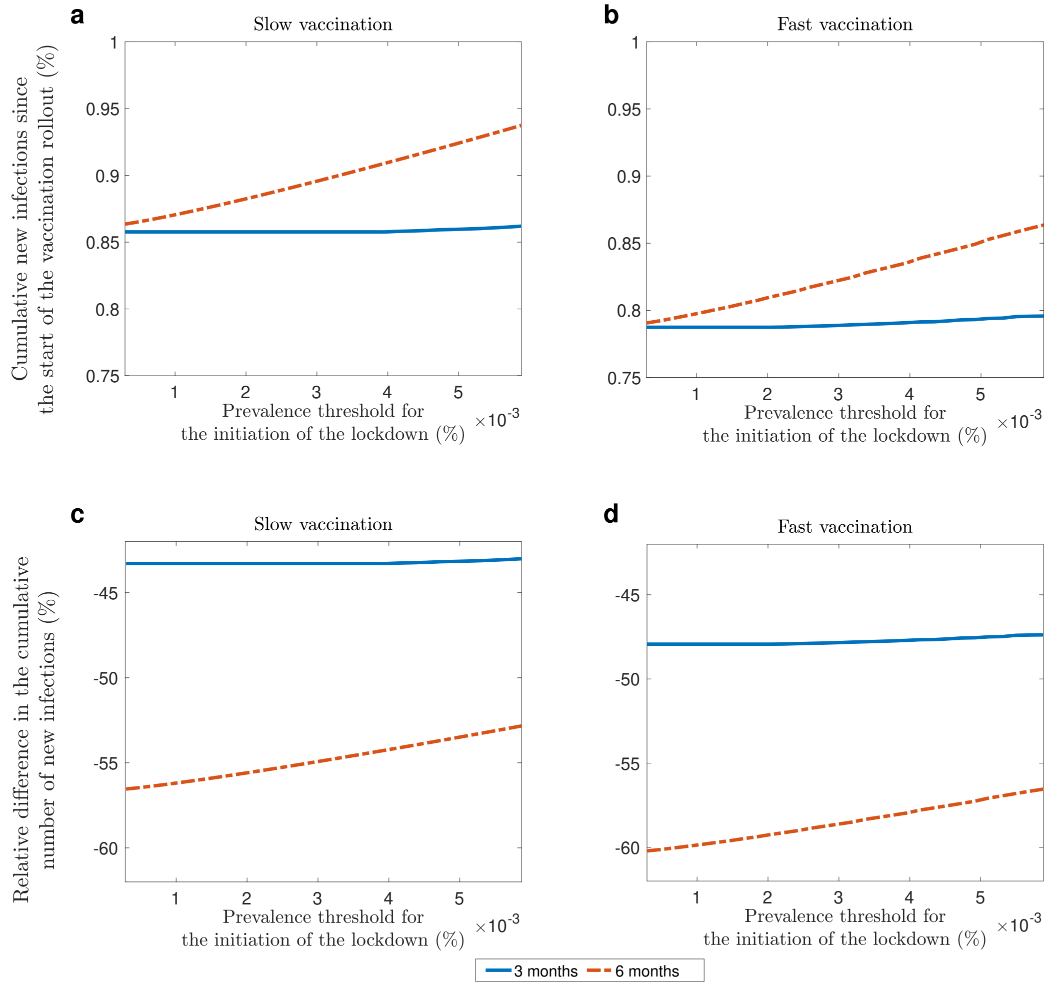

Figure 3: **Cumulative number of new infections for different thresholds of the initiation of lockdown restriction.** **a** and **b** show the cumulative number of new infections, presented as a percentage of the total population size. **c** and **d** show the relative difference in the number of new infections as compared to the no-vaccination scenario. **a** and **c** show the outputs for the slow vaccination uptake, **b** and **d** for the fast vaccination uptake.

We also investigated the improvements achieved by supplementing the vaccination rollout with a "dynamics" lockdown (Figure 4-5). We observe that the "dynamic" lockdown can lower the cumulative number of new infections almost two fold in the short term (three months after the start of the vaccination rollout) and more than that in the long term (six months after the start of the vaccination rollout) as compared with no-vaccination scenario. Supplementing the vaccination rollout with this intervention yields the best improvements on the no-vaccination scenario for a fast vaccination rate and a vaccine with high efficacy. On the other hand, when comparing the vaccination rollout with "dynamic" lockdown to one without, we observed that the largest improvements are gained for a fast vaccination rate and a vaccine with low efficacy. The lowest improvement are gained for a slow vaccination rate and a vaccine with high efficacy.

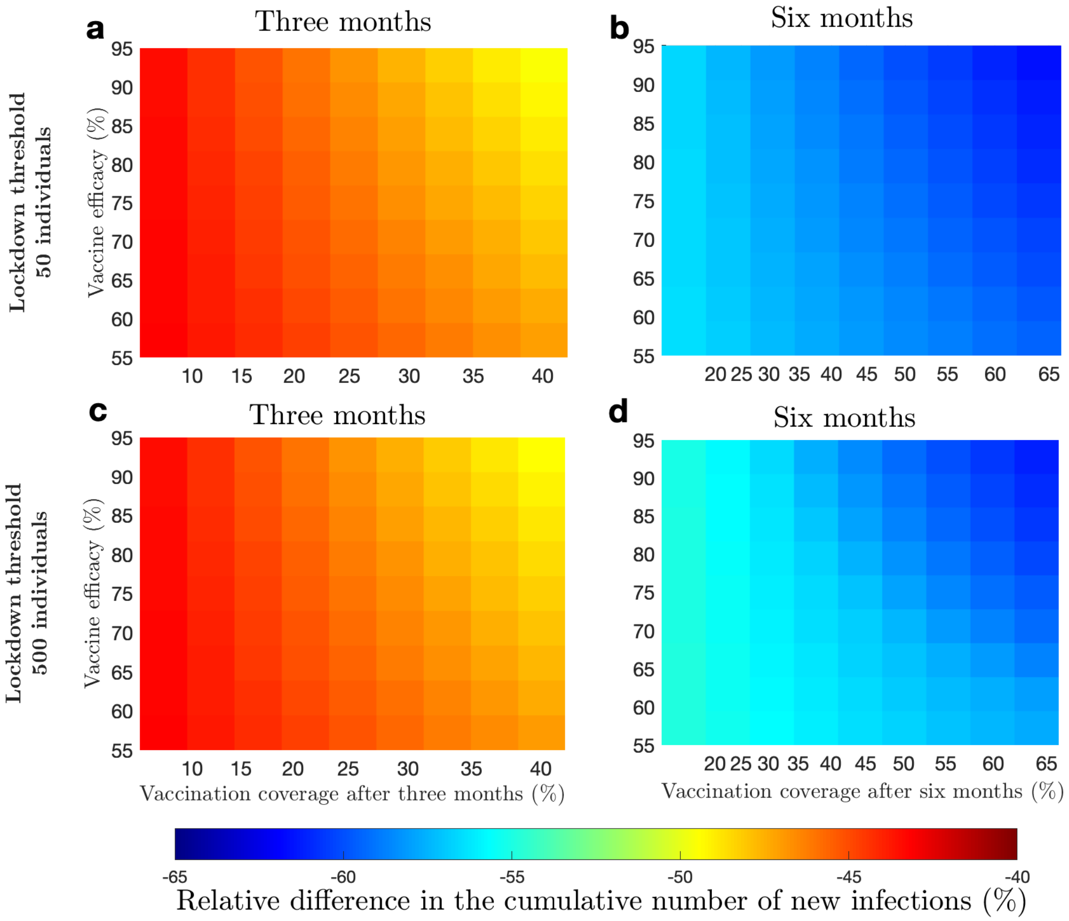

Figure 4: **Relative difference in the cumulative number of new infections as compared to the no-vaccination scenario for different thresholds of initiation of lockdown strengthening.** Difference in the cumulative number of new infections as compared to the no-vaccination scenario after **a** and **c** three months; **b** and **d** six months of the vaccination rollout. **a** and **b** Results for a lockdown threshold of 50 individuals, **c** and **d** for a lockdown threshold of 500 individuals.

### 3 Sensitivity analyses

In this section we report results on the sensitivity of the epidemic dynamics during vaccination rollout to assumptions about initial conditions and parameter values. We considered the cumulative number of new infections three and six months after the start of the vaccination rollout. We used the absolute size of the cumulative number, presented as percentage of the total population size and the relative difference with respect to the cumulative number of new infections relative to the no-vaccination scenario, presented as percentage. The original variant of the virus circulates and no interventions targeting compliance are in place.

#### 3.1 Initial conditions

First, we investigated sensitivity of the results to the initial sizes of the compartments at the start of the simulation. More specifically, we varied the initial numbers of the compliant, exposed, infectious, and recovered populations in the ranges of 20-90%, 0.1-1% 0.01-1%, and 5-20%, respectively. The model parameters were fixed to the values

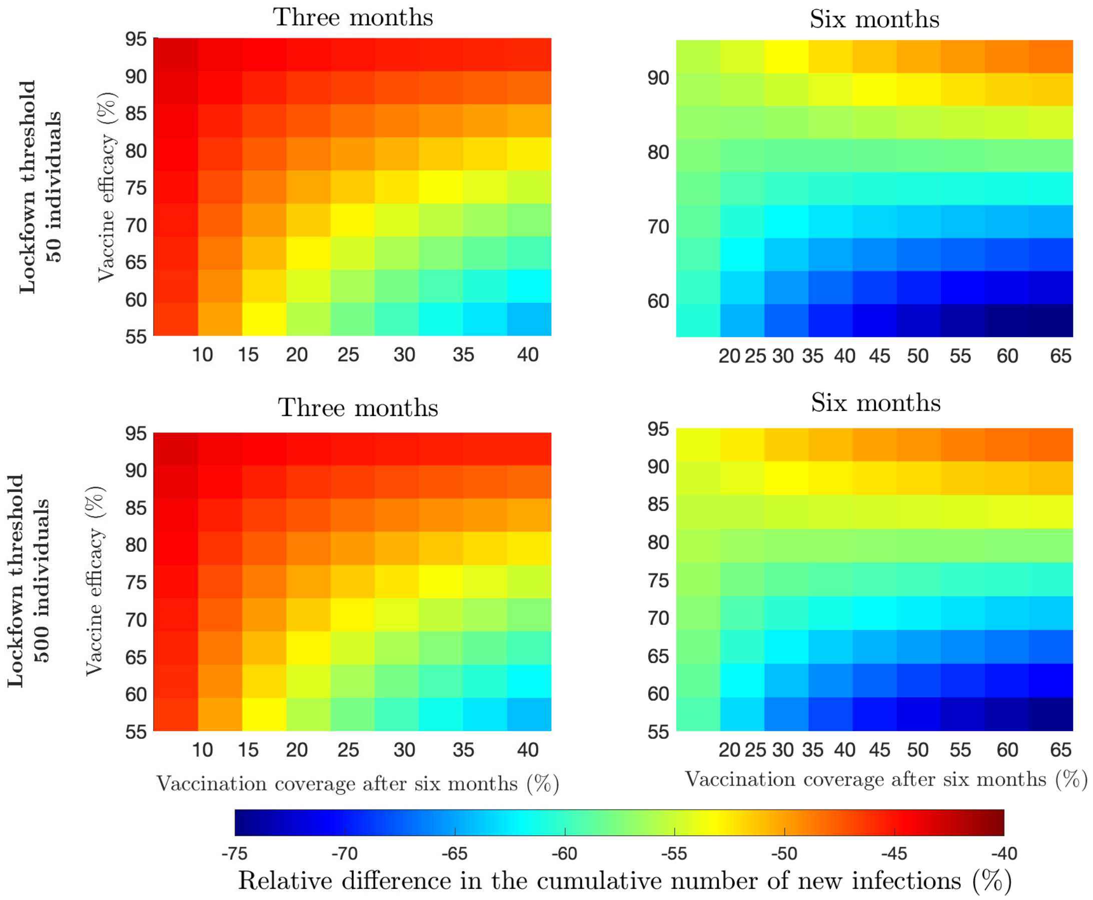

Figure 5: **Relative difference in the cumulative number of new infections as compared to the vaccination rollout without additional interventions for different thresholds of initiation of lockdown strengthening.** Difference in the cumulative number of new infections as compared to the vaccination rollout without additional interventions after **a** and **c** three months; **b** and **d** six months of the vaccination rollout. **a** and **b** Results for a lockdown threshold of 50 individuals, **c** and **d** for a lockdown threshold of 500 individuals.

used in the main text, with vaccine efficacy in preventing the acquisition of the infection set at 60%. The results are presented for slow and fast vaccination rates (see the main text for the definition).

##### 3.1.1 Compliant proportion of the population

In the main analysis, we calibrated the percentage of the population compliant with physical distancing measures at the start of the vaccination rollout using reported compliance of 65% with a specific measure (keeping 1.5m distance) in the Netherlands on the week of November 11-17, 2020 [2]. We used this number as a proxy to being compliant to recommended physical distancing measures, and subsequently substantially reducing contact rates. In what follows, we vary the initial percentage in a range of $20-90\%$ for the percentage of the population that complies with physical distancing measures, and investigate the effect of the initial percentage of compliance on the outputs (Figure 6). The sizes of susceptible, exposed, infectious and recovered compartments are fixed to the values used in the main analysis.

The model predicts that the cumulative number of new infections is lower for higher percentage of the initial

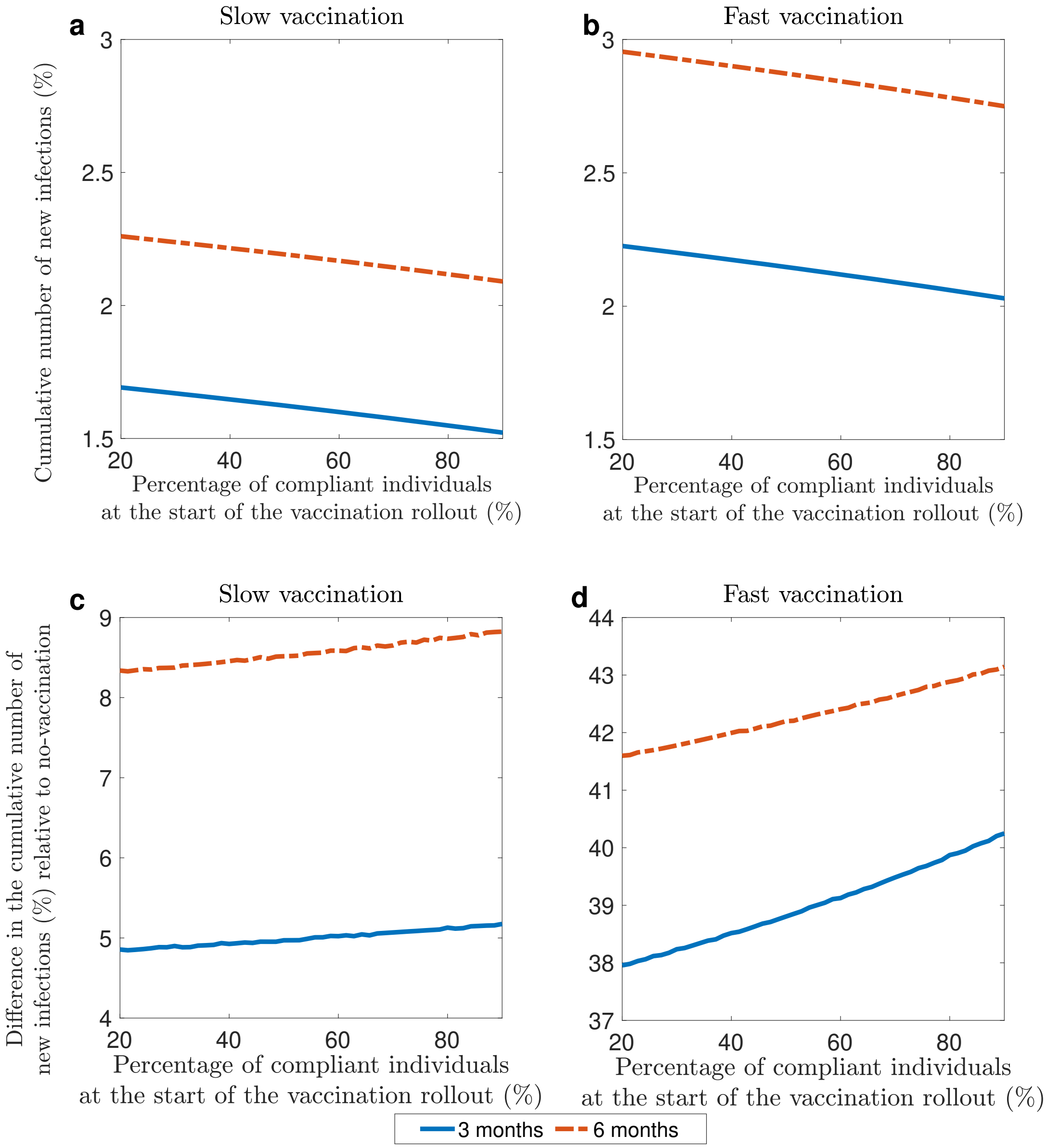

Figure 6: **Cumulative number of new infections for different percentages of compliant individuals at the start of the vaccination rollout. a** and **b** show cumulative number of new infections versus percentage of compliant individuals at the start of the vaccination rollout. The results are presented as a percentage of the total population. **c** and **d** show relative difference in the cumulative number of new infections relative to the baseline no-vaccination values versus percentage of compliant individuals at the start of the vaccination rollout. The results are presented as a percentage of the cumulative number of new infections in the no-vaccination scenario. The original variant is circulating. The results are presented for slow (**a** and **c**) and fast (**b** and **d**) vaccination rates.

proportion of compliant individuals. This is observed in the short term (three months following the vaccination rollout, Figure 6**a**) and in the long term (six months following the vaccination rollout, Figure 6**b**).

The model predicts that the excess of infections reported in the main analysis is preserved for the range of percentages of compliant individuals that we considered (Figure 6**c** and 6**d**). This percentage is an increases as the initial proportion of compliant individuals increases and is higher for a fast vaccination rollout following three and six of the vaccination rollout. However, variation of relative excess of the infections as the percentage of compliant individuals change does not exceed 3%. This indicates the outputs are not sensitive to the variation in the initial number of compliant individuals.

##### 3.1.2 Seroprevalence

We defined seroprevalence as the proportion of the population that has been infected with SARS-CoV-2 and is immune to a new infection at the start of the simulations. In the main analysis we calibrated the model to a seroprevalence of 8%, which is between what was measured in the Netherlands in September/October 2020 [3] and in February 2021 [4]. We explored the sensitivity of the outputs to the initial value of seroprevalence, by varying the initial seroprevalence in the range of 5-20% (Figure 7). We kept the sizes of the exposed and infectious compartments fixed to the values used in the main analysis. To preserve the constant size of the total population, we adjusted the size of the susceptible compartment accordingly.

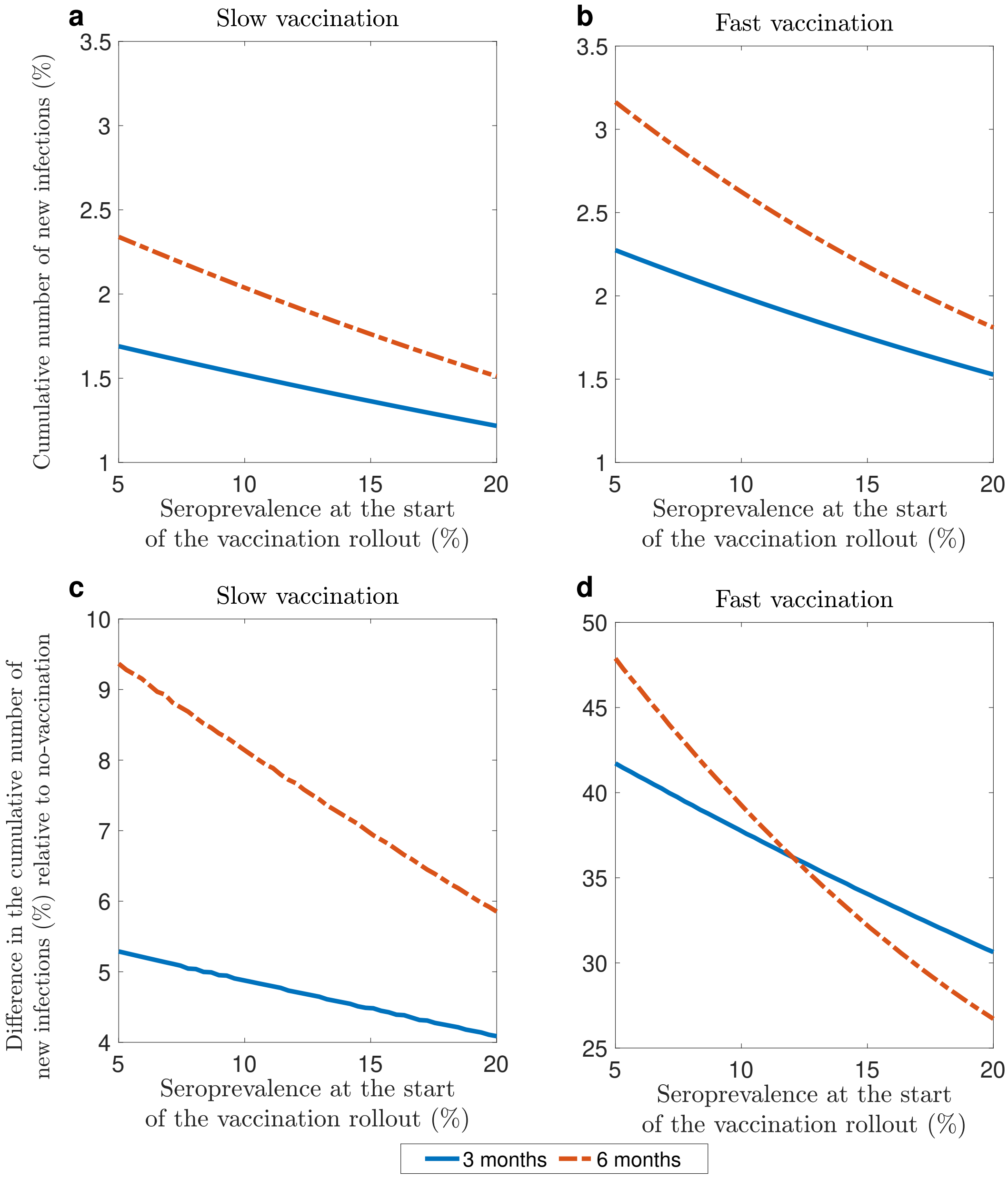

Figure 7: **Cumulative number of new infections for different seroprevalence at the start of the vaccination rollout.** **a** and **b** show the cumulative number of new infections versus percentage of recovered individuals at the start of the vaccination rollout. The results are presented as a percentage of the total population. **c** and **d** show relative difference in the cumulative number of new infections relative to the baseline no-vaccination values versus percentage of recovered individuals at the start of the vaccination rollout. The results are presented as a percentage of the cumulative number of new infections in the no-vaccination scenario. The original variant is circulating. The results are presented for slow (**a** and **c**) and fast (**b** and **d**) vaccination rates.

The model predicts that the cumulative number of new infections is lower for higher seroprevalence. This is observed in the short term (three months following the vaccination rollout, Figure 7**a**) and in the long term (six months following the vaccination rollout, Figure 7**b**).

Our simulations show that the excess infections seen in the main analysis is preserved for a wide range of seroprevalence values (Figure 7**c** and 7**d**). For a fast vaccination rate the relative excess is significantly larger than for a slow vaccination rate, both in the long and in the short term. For both slow and fast vaccination, the relative excess of infections is decreasing as the percentage of recovered individuals at the start of the vaccination rollout increases. Noteworthy, this decrease is much faster for the fast vaccination rollout than for the slow one, making the dynamics very sensitive to the value of seroprevalence at the start of the vaccination rollout.

##### 3.1.3 Proportion of infectious cases

In the main analysis we set the number of infectious individuals to be equal to 112,435 individuals (0.66% of the population size of the Netherlands) as was estimated by RIVM for the week November 11-17. We explored the sensitivity of the outputs to the initial value of the number of infectious cases, which we sampled from the interval 0.1-1% (Figures 8). We kept the sizes of the exposed and recovered compartments fixed to the values used in the main analysis. To preserve the constant size of the total population, we adjusted the size of the susceptible compartment accordingly.

The model predicts that the cumulative number of new infections increases as the number of infectious individuals at the start of the vaccination rollout increases. This is observed in the short term (three months following the vaccination rollout, Figure 8**a**) and in the long term (six months following the vaccination rollout, Figure 8**b**).

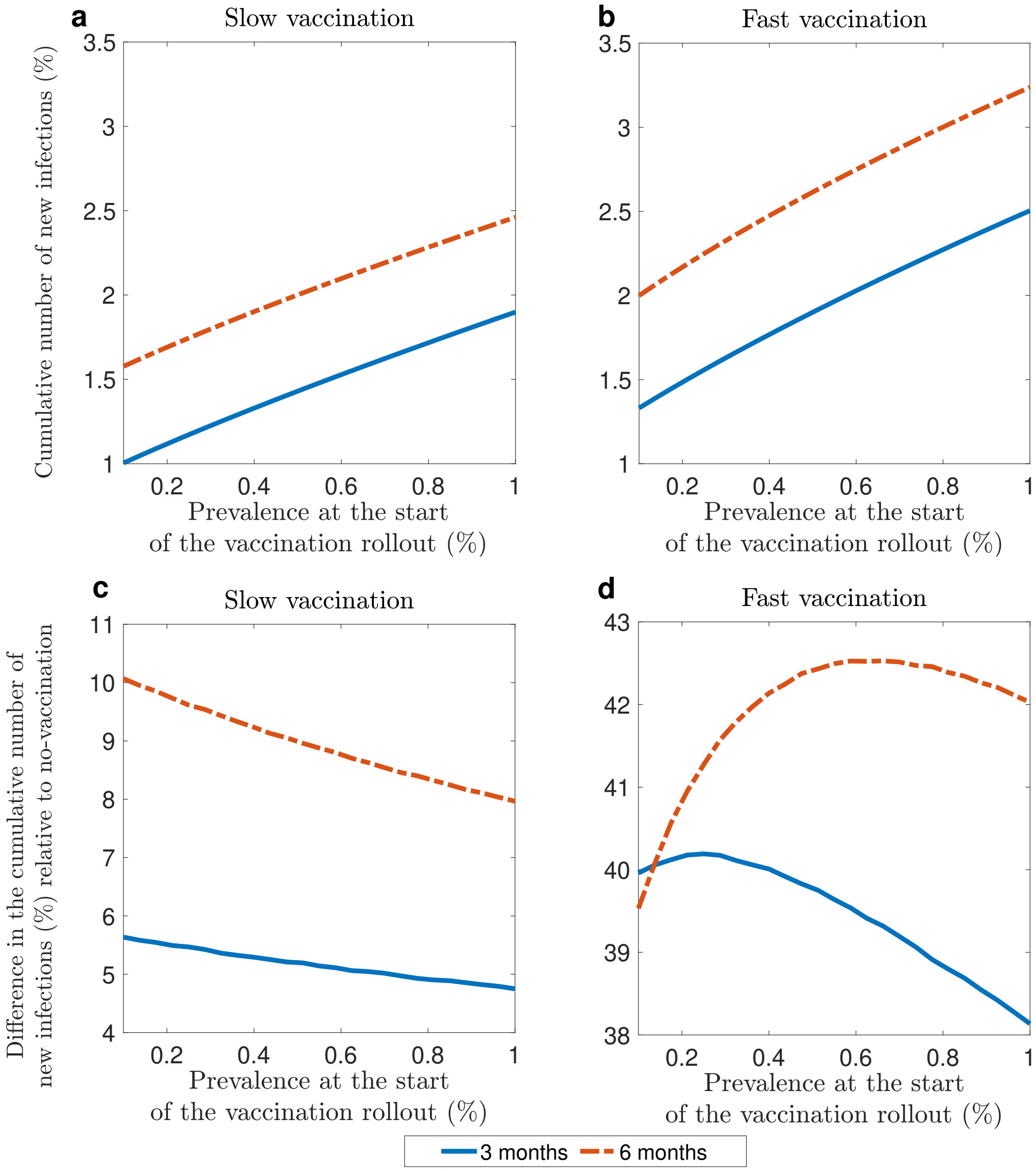

Figure 8: **Cumulative number of new infections for different percentages of infectious individuals at the start of the vaccination rollout.** **a** and **b** show the cumulative number of new infections versus percentage of recovered individuals at the start of the vaccination rollout. The results are presented as a percentage of the total population. **c** and **d** show relative difference in the cumulative number of new infections relative to the baseline no-vaccination values versus percentage of infectious individuals at the start of the vaccination rollout. The results are presented as a percentage of the cumulative number of new infections in the no-vaccination scenario. The original variant is circulating. The results are presented for slow (**a** and **c**) and fast (**b** and **d**) vaccination rates.

Our simulations show that the excess infections seen in the main analysis is preserved for a wide range of initial infectious individuals values (Figure 8**c** and 8**d**). For slow vaccination rollout, the excess is decreasing with increasing number of infectious individuals, both in the long term and in the short term. In contrast, for a fast vaccination rate, for a low initial initial number of infectious individuals, the excess increases, while for a higher number it decreases. This relationship is present both in the short term (three months after the start of the vaccination rollout) and in the long term (six months after the start of the vaccination rollout). We note that changes in the relative excess of infections in the range of the number of infectious individuals that we considered does not exceed 3%, thus indicating a low sensitivity of the outputs to variations in this initial condition.

###### 3.1.4 Proportion of exposed cases

In the main analysis we set number of infectious of exposed individuals to be equal to 64249 individuals (0.38% of the population size of the Netherlands) which we calculate using the approximation to the total number of infectious cases made by RIVM for the week November 11-1. We explored the impact of the initial proportion of exposed cases on epidemic and compliance dynamics by sampling the prevalence in the range of 0.1-1% of the total population (Figures 8). As the size of the exposed compartment changed, we kept the size of the infectious and recovered compartments fixed to the values used in the main analysis. To preserve the constant size of the total population, we adjusted the size of the susceptible compartment.

The model predicts that the cumulative number of new infections increases as the proportion of exposed cases at the start of the vaccination rollout increases. This is observed in the short term (three months following the vaccination rollout, Figure 9**a**) and in the long term (six months following the vaccination rollout, Figure 9**b**).

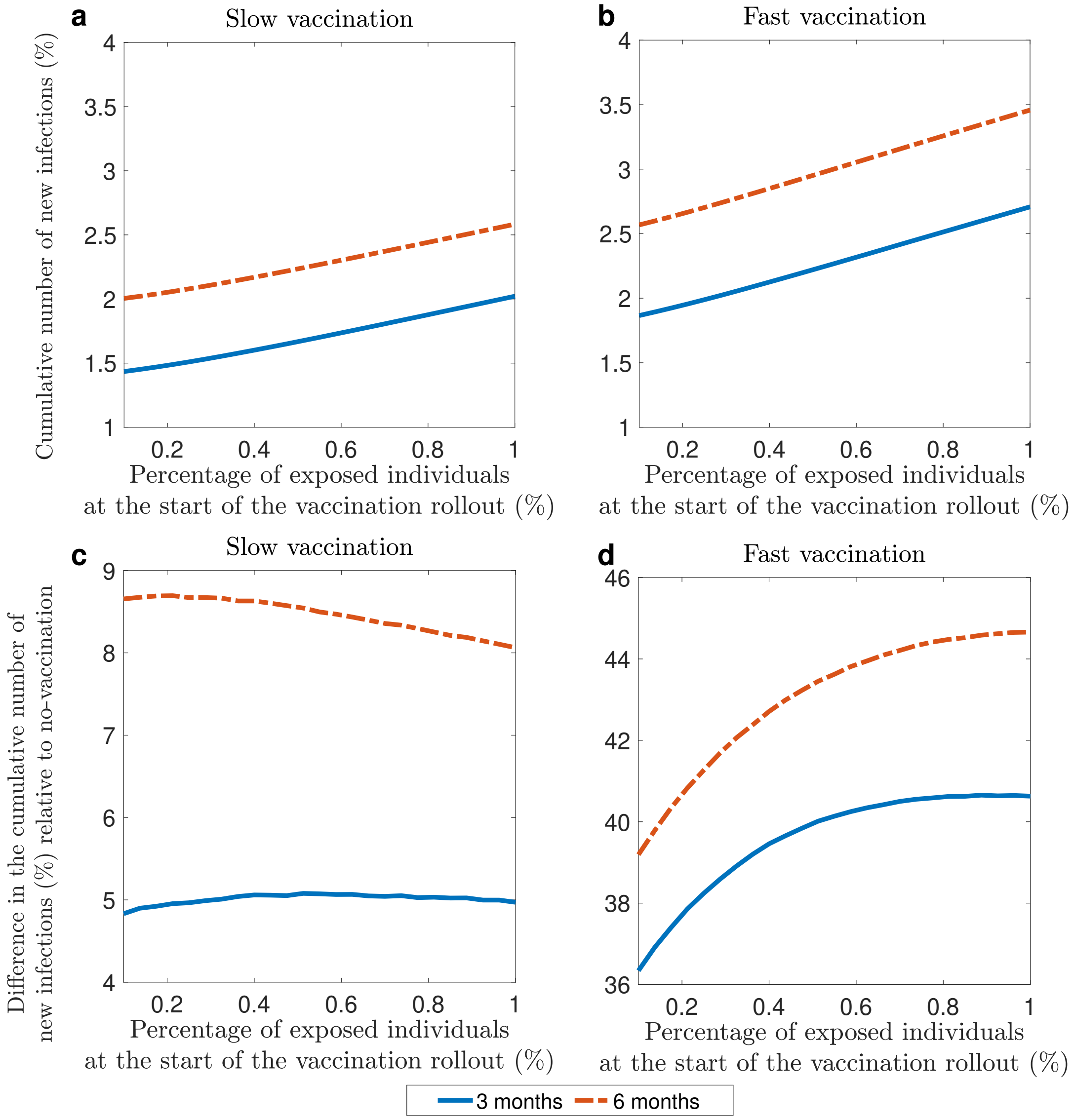

Figure 9: **Cumulative number of new infections for different percentages of exposed individuals at the start of the vaccination rollout.** **a** and **b** show the cumulative number of new infections versus percentage of recovered individuals at the start of the vaccination rollout. The results are presented as a percentage of the total population. **c** and **d** show relative difference in the cumulative number of new infections relative to the baseline no-vaccination values versus percentage of exposed individuals at the start of the vaccination rollout. The results are presented as a percentage of the cumulative number of new infections in the no-vaccination scenario. The original variant is circulating. The results are presented for slow (**a** and **c**) and fast (**b** and **d**) vaccination rates.

Our simulations indicate that the excess of the new infections as compared to the baseline no-vaccination scenario is preserved for all the values of percentage of exposed individuals that we have sampled. We also observe a relatively low sensitivity of the relative excess of new infections to changes in the initial percentage of exposed individuals (Figures 9**c** and 9**d**) when the vaccination uptake is low. In this case, the relative excess of the cumulative number of infections remains on approximately the same level on the whole range that we considered. On the other hand, given the fast vaccination rate, we observe that the relative excess increases as the initial proportion of exposed individuals increases and that the outputs corresponding to endpoints of the exposed percentage interval are approximately 5%

apart, both for three and six months.

#### 3.2 Sensitivity analysis with respect to model parameters

In this section we report results of the investigation of sensitivity of the outputs of the model to the chosen values of parameters. The outputs are the cumulative number of new infections three and six months after the vaccination rollout started presented as the percentage from the total population size. The initial conditions are fixed to the values that were used in the main text. The parameters that we consider are 1. the duration of the exposed period ($1/\alpha$); 2. the duration of the infectious period ($1/\gamma$); 3. the average contact rate of non-compliant individuals ($c$); 4. the average contact rate of compliant individuals ($cr_1$); 5. rate of moving to compliant state ($\delta$); 6. the duration of compliant state when there is no vaccination ($\mu_0$). We look at the effects of variation parameters in pairs, fixing the rest of the parameters to be equal to the values used in the main analysis. Similarly, the initial conditions are fixed to be equal to the values used in the main analysis. The results are presented for slow and fast vaccination rates (see the main text for the definition).

##### 3.2.1 Duration of latent and infectious periods

In this section we consider the sensitivity of the outputs to the selected values of the duration of the exposed period ($1/\alpha$) and the duration of the infectious period ($1/\gamma$). In the main text they are fixed to be 4 and 7 days, respectively. Here we sample $1/\alpha$ in the range of 2-6 days and $1/\gamma$ in the range of 5-9 days (Figure 10).

We observe that the epidemic burden increases as the infectious period increases, such that when the vaccination rate is fast the increase in the cumulative number of new infections is higher than when the vaccination rate is slow. On the other hand, we observe that when the length of the exposed period has very little bearing on the cumulative number of new infections, as compared to the duration of infectious period.

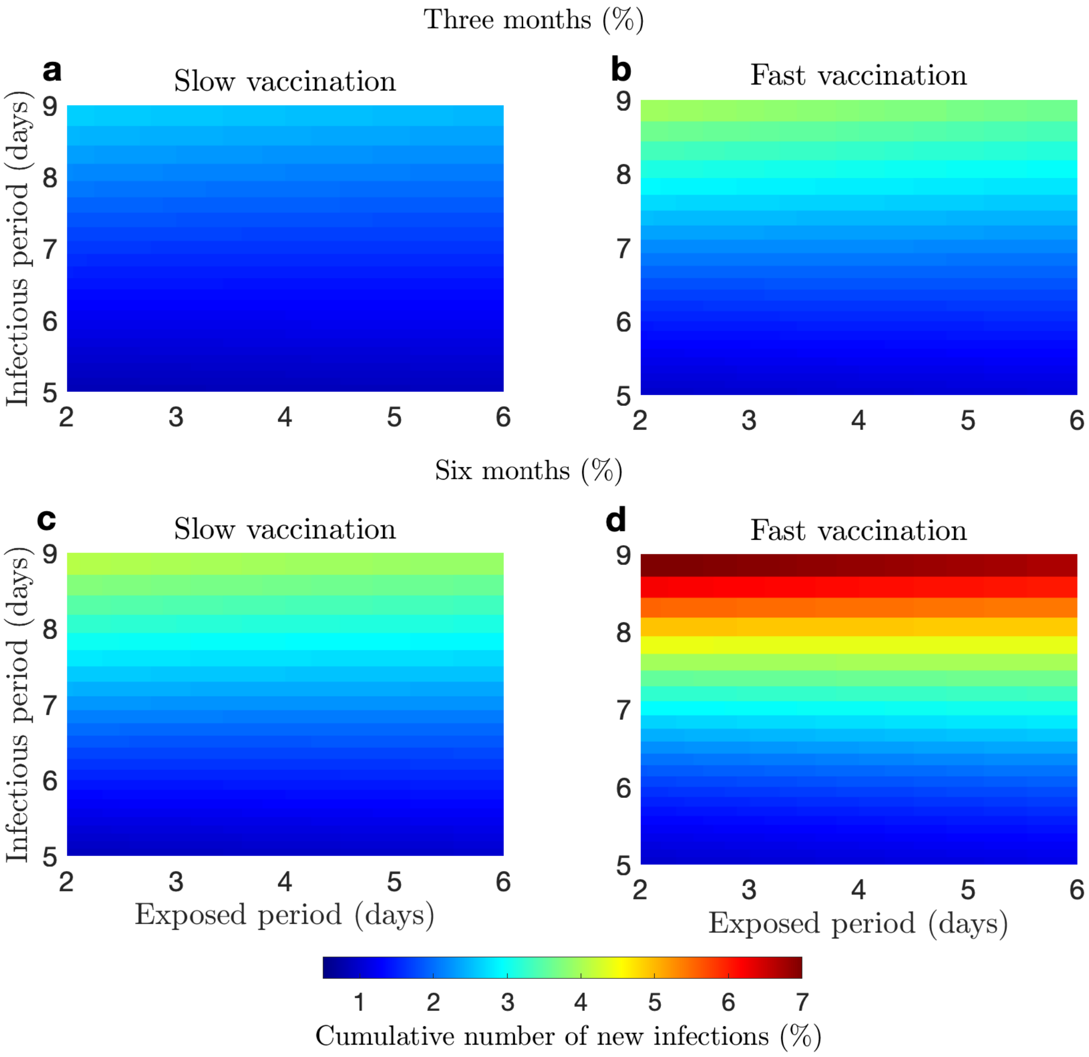

Figure 10: **Cumulative number of new infections depending on duration of exposed and infectious periods.** **a** and **b** show the cumulative number of infections three months after the start of vaccination rollout. **c** and **d** show the cumulative number of infections six months after the start of vaccination rollout. **a** and **c** show these quantities for the slow vaccination uptake, **b** and **d** for the fast vaccination uptake.

Our results indicate the relative excess of infections as compared to the no-vaccination scenario is preserved throughout the ranges that we have considered (Figure 11). However, the sensitivity of the magnitude of the excess to variation in the duration of exposed and infectious periods depend on the vaccination uptake rate. If the vaccination rate is slow, than the largest change in the excess that we have measured across the parameter range was approximately equal to 13%. For the vast vaccination rate, especially at a later time the expected excess ranged from almost 14% to 99%. The excess in the cumulative number of infections is increasing as either the duration of the infectious period and of the duration of the exposed period increases. However, the changes are more drastic for the former than for the latter.

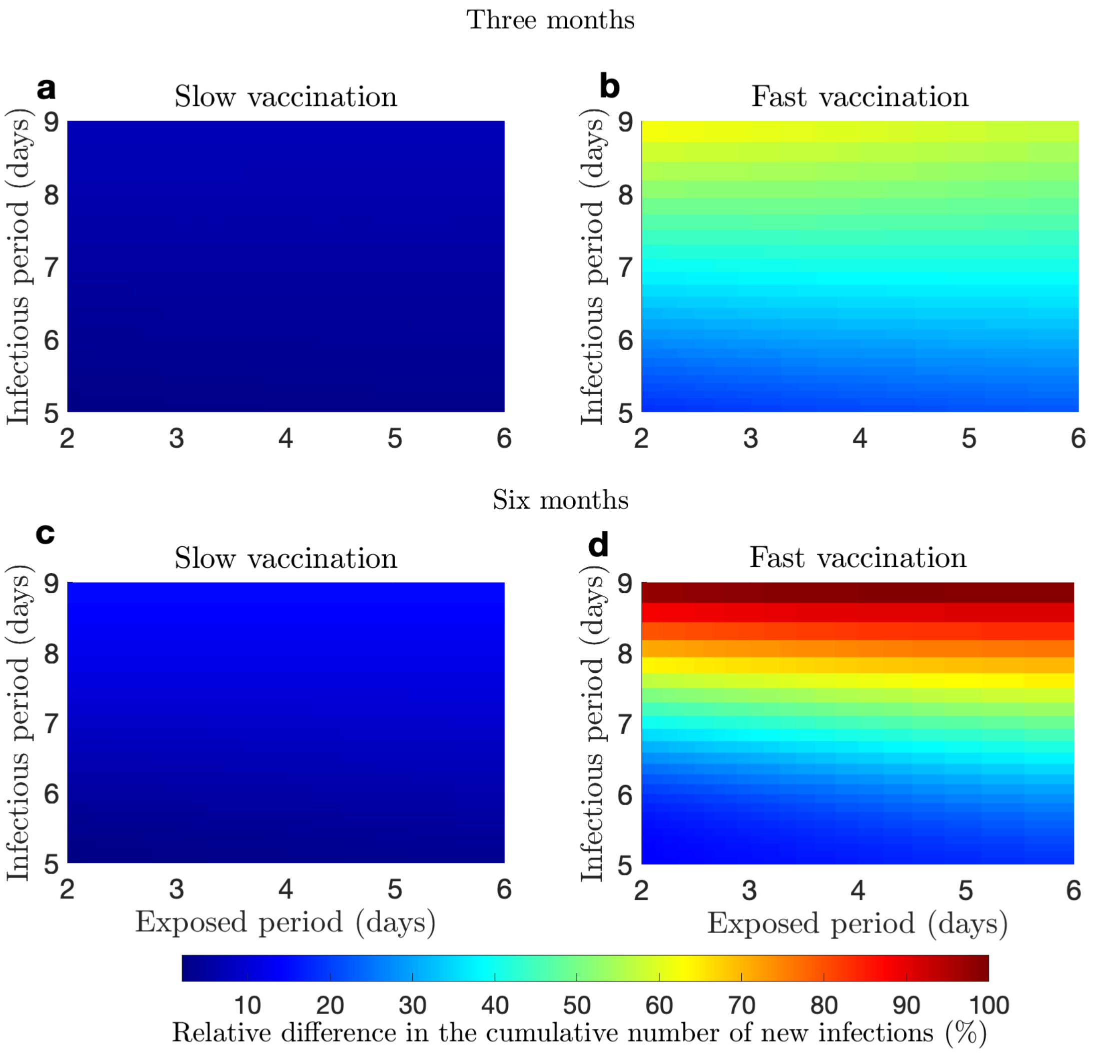

Figure 11: **Relative difference in the cumulative number of new infections compared to the no-vaccination scenario depending on duration of exposed and infectious periods.** **a** and **b** show relative difference in the cumulative number of new infections three months after the start of vaccination rollout; **c** and **d** show the same quantity six months after the start of vaccination rollout. **a** and **c** slow vaccination uptake; **b** and **d** fast vaccination uptake.

##### 3.2.2 Contact rates of compliant and non-compliant individuals

We considered the sensitivity of the outputs to the contact rates of compliant individuals $c$ and the ratio of contact rates of compliant and non-compliant individuals $r_1$. In the main text these parameters were fixed at 8.8 per day and 0.34, respectively. Here we vary $c$ in the range of 0.5-15 per day and $r_1$ in the range of 0.01-1 (Figure 12). The effective reproduction number changes as $c$ and $r_1$ change.

We observe that both parameters have a strong influence on the cumulative number of infections, both in the short term (after three months of the vaccination rollout) and in the long term (after six months of the vaccination rollout). At both these time points, fast vaccination is characterized by significantly lower cumulative number of new infections, both in the short term and in the long term.

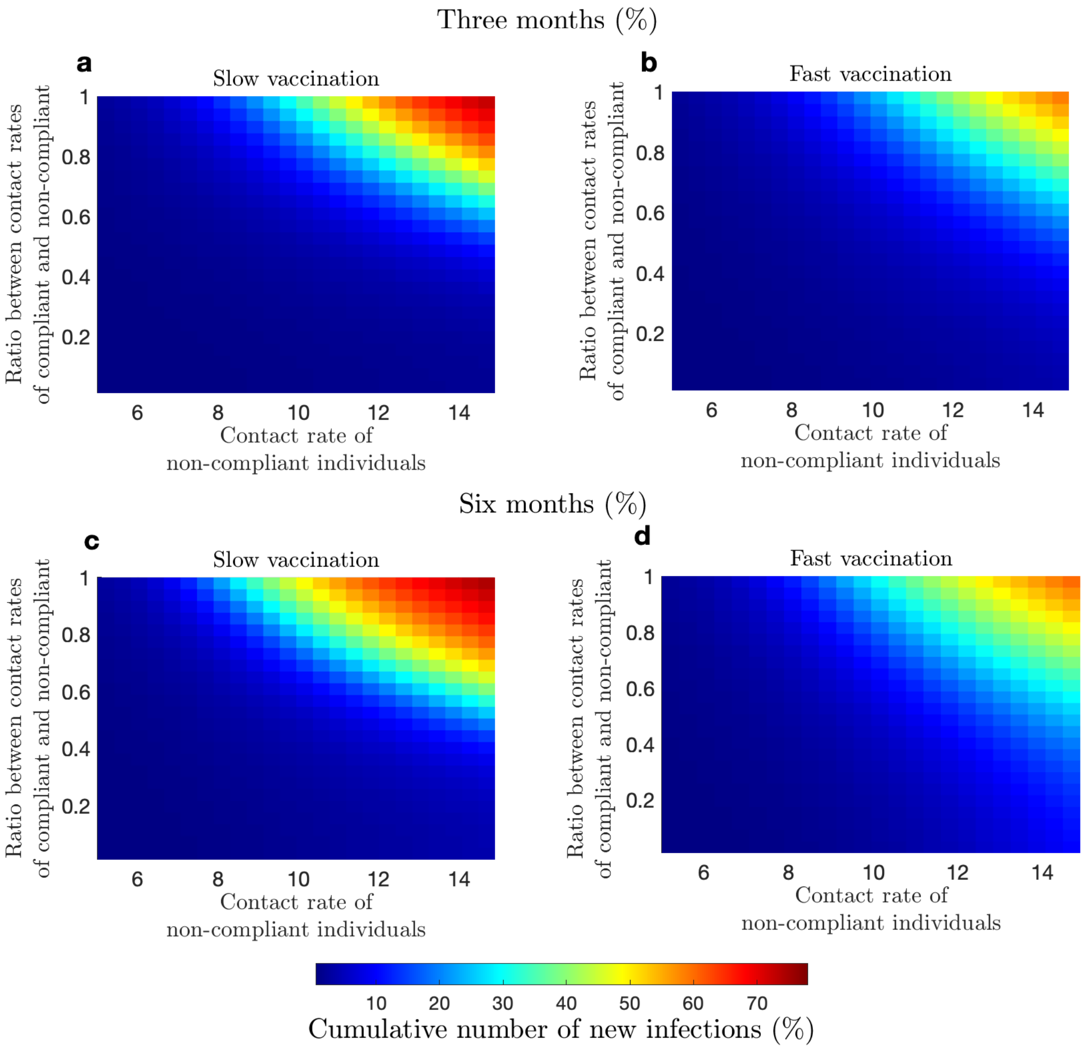

Figure 12: **Cumulative number of new infections for different depending on contact rates of compliant and non-compliant individuals.** **a** and **b** show the cumulative number of infections three months after the start of vaccination rollout; **c** and **d** six months after the start of vaccination rollout. **a** and **c** show these quantities for the slow vaccination uptake, **b** and **d** fast vaccination uptake.

Our simulations shown in Figure 13 indicate that a possible excess in number of infections as compared to the no-vaccination scenario is highly sensitive to the contact rates of compliant ad non-compliant individuals. Generally, we expect the cumulative number of infections to exceed that of the no-vaccination scenario if there is a significant difference between contact rates of compliant and non-compliant individuals. The largest increases in the cumulative number of infections in the first months of the vaccination rollout are expected when the contact rate of non-compliant individuals is close to the pre-pandemic levels and the contact rate of compliant individuals is significantly lower.

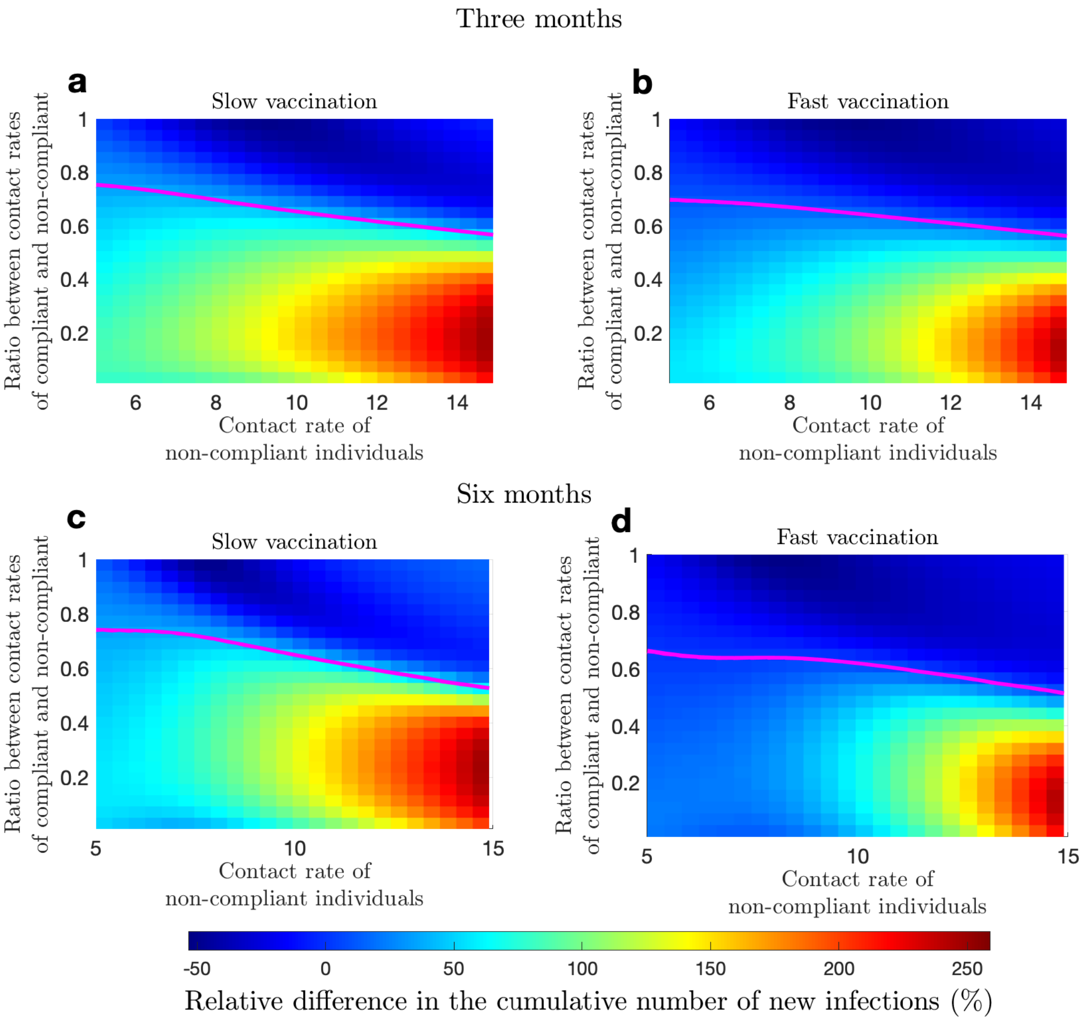

Figure 13: **Relative difference in the cumulative number of new infections compared to the no-vaccination scenario depending on contact rates of compliant and non-compliant individuals.** **a** and **b** show the relative difference in the cumulative number of new infections compared to the no-vaccination scenario three months after the start of vaccination rollout; **c** and **d** show the same quantity six months after the start of vaccination rollout. **a** and **c** show these quantities for the slow vaccination uptake, **b** and **d** fast vaccination uptake.

##### 3.2.3 Compliance acquisition and loss rates

We considered the sensitivity of the outputs to the rate of moving to the compliant state ($\delta$), and to the duration of compliance when there is no vaccination ($1/mu_0$). In the main text we set the compliance duration when there is no vaccination to 30 days. This is an assumed value and here we test the effect of shorter duration of compliance on epidemic dynamics. We consider a range of compliance duration between 7 and 30 days. In the main text we fixed the rate of moving to the compliant state to $4 \times 10^{-5}$ per day. Here, we considered the range of $10^{-6}$-$10^{-4}$. The results are summarized in Figure 14

We observe that the outputs are sensitive to the values of both parameters with the cumulative number of infections decreasing as the rate of moving to compliant state increases and duration of compliant state. However, this relationship is more apparent six month after the start of the vaccination rollout than after three months.

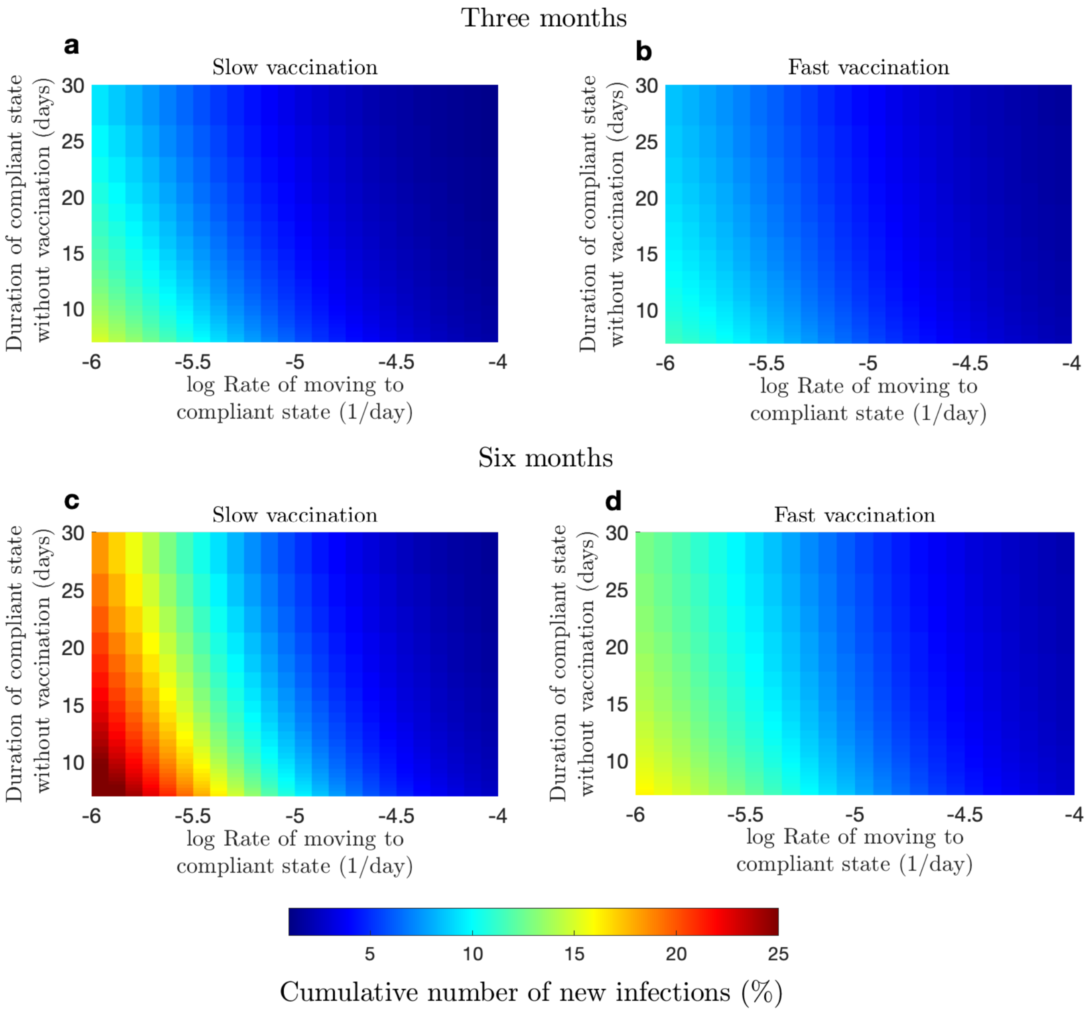

Figure 14: **Cumulative number of new infections depending on the rate of moving to the compliant state and compliance duration.** **a** and **b** show the cumulative number of infections three months after the start of vaccination rollout; **c** and **d** six months after the start of vaccination rollout. **a** and **c** show these quantities for the slow vaccination uptake, **b** and **d** fast vaccination uptake.

Our simulations indicate that the occurrence of excess infections relative to the no-vaccination scenario during the first months of the vaccination rollout is sensitive to changes in the rate of moving to the compliant state and the duration of the compliant state after the first three months of vaccination. The excess of infections is observed for high rates of moving to the compliant state and long duration of compliance. After six months of vaccination there was an excess of infections for the whole range of parameters that we considered.

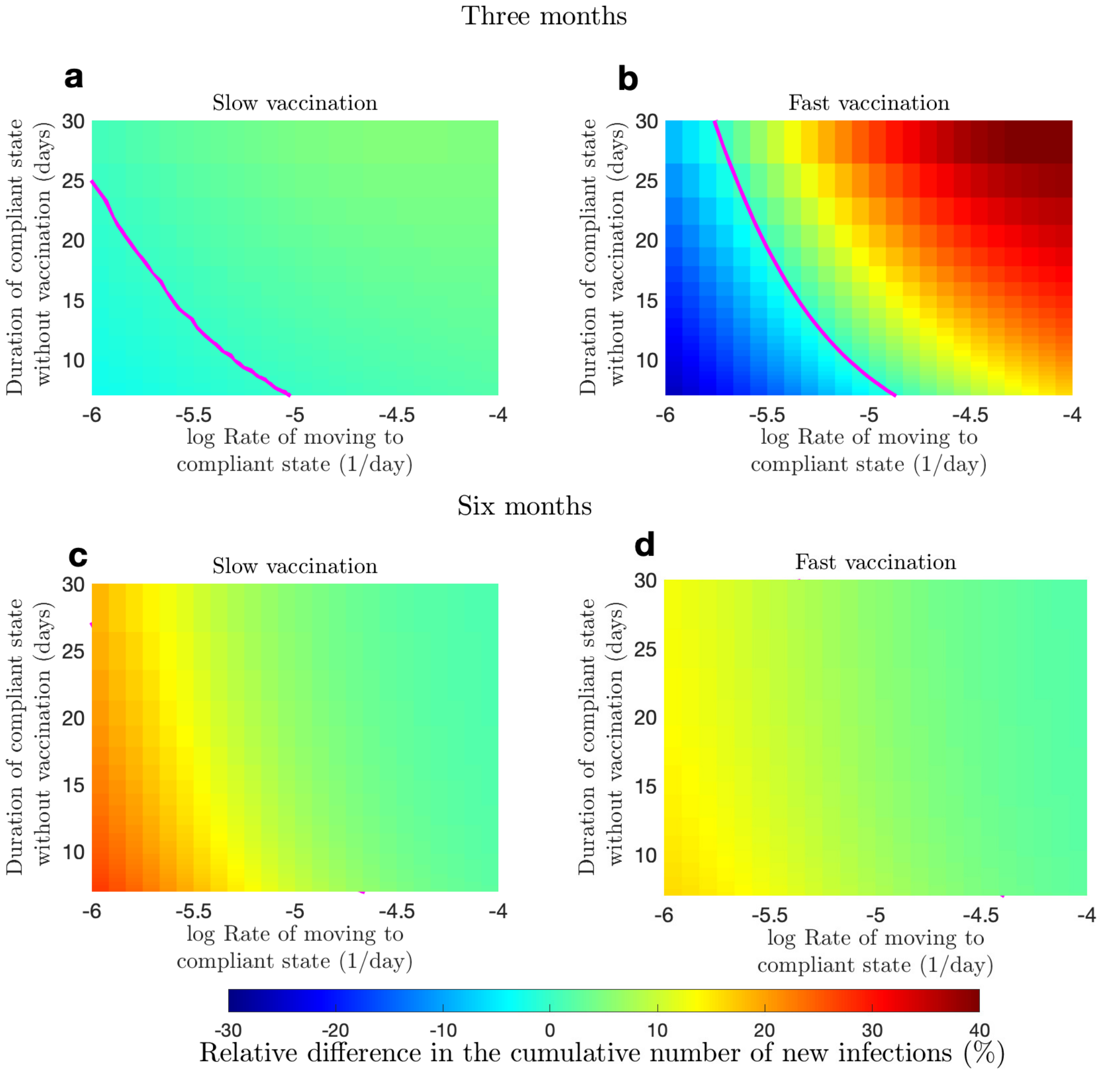

Figure 15: **Relative difference in the cumulative number of new infections compared to the no-vaccination scenario depending on transition rate to compliance and compliance duration.** **a** and **b** show the relative difference in the cumulative number of new infections compared to the no-vaccination scenario three months after the start of vaccination rollout; **c** and **d** six months after the start of vaccination rollout. **a** and **c** show these quantities for the slow vaccination uptake, **b** and **d** fast vaccination uptake.
